# Cluster analysis of transcriptomic datasets to identify endotypes of Idiopathic Pulmonary Fibrosis

**DOI:** 10.1101/2021.07.16.21260633

**Authors:** Luke M Kraven, Adam R. Taylor, Philip L. Molyneaux, Toby M. Maher, John E. McDonough, Marco Mura, Ivana V. Yang, David A. Shwartz, Yong Huang, Imre Noth, Shwu-Fan Ma, Astrid J. Yeo, William A. Fahy, R. Gisli Jenkins, Louise V. Wain

## Abstract

**Background:** Considerable clinical heterogeneity in Idiopathic Pulmonary Fibrosis (IPF) suggests the existence of multiple disease endotypes. Identifying these endotypes could allow for a biomarker-driven personalised medicine approach in IPF. To improve our understanding of the pathogenesis of IPF by identifying clinically distinct groups of patients with IPF that could represent distinct disease endotypes.

**Methods:** We co-normalised, pooled and clustered three publicly available blood transcriptomic datasets (total 220 IPF cases). We compared clinical traits across clusters and used gene enrichment analysis to identify biological pathways and processes that were over-represented among the genes that were differentially expressed across clusters. A gene-based classifier was developed and validated using three additional independent datasets (total 194 IPF cases).

**Findings:** We identified three clusters of IPF patients with statistically significant differences in lung function (P=0·009) and mortality (P=0·009) between groups. Gene enrichment analysis implicated dysregulation of mitochondrial homeostasis, apoptosis, cell cycle and innate and adaptive immunity in the pathogenesis underlying these groups. We developed and validated a 13-gene cluster classifier that predicted mortality in IPF (high-risk clusters vs low-risk cluster: hazard ratio= 4·25, 95% confidence interval= [2·14, 8·46], P=3·7×10^−5^).

**Interpretation:** We have identified blood gene expression signatures capable of discerning groups of IPF patients with significant differences in survival. These clusters could be representative of distinct pathophysiological states, which would support the theory of multiple endotypes of IPF. Although more work must be done to confirm the existence of these endotypes, our classifier could be a useful tool in patient stratification and outcome prediction in IPF.

**Funding:** L.V.W. holds a GSK/British Lung Foundation Chair in Respiratory Research (C17-1). R.G.J. is supported by a National Institute for Health Research (NIHR) Research Professorship (NIHR reference RP-2017-08-ST2-014). P.L.M. is supported by an Action for Pulmonary Fibrosis Mike Bray fellowship. T.M. Maher is supported by a National Institute for Health Research Clinician Scientist Fellowship (CS-2013-13-017) and a British Lung Foundation Chair in Respiratory Research (C17-3). I.N. is supported by a National Heart, Lung, and Blood Institute (NHLBI) grant (R01HL145266). D.A.S. is supported by NHLBI grants (UG3HL151865, R01HL097163, P01HL092870, X01HL134585 and UH3HL123442) and a United States Department of Defense grant (W81XWH-17-1-0597). The GSE110147 study was supported by the Roche Multi Organ Transplant Academic Enrichment Fund, Lawson Research Institute Internal Research Fund and Western Strategic Support for CIHR Success, Seed Grant. The research was partially supported by the NIHR Leicester Biomedical Research Centre; the views expressed are those of the author(s) and not necessarily those of the National Health Service (NHS), the NIHR, or the Department of Health.

**Putting research into context:** *Evidence before this study:* We searched PubMed Central in February 2020 with the search terms “idiopathic pulmonary fibrosis”, “gene expression” and “cluster analysis” with no restrictions on publication date or language. Previous transcriptomic cluster analyses have found that differences in gene expression can be used to predict disease status, severity and outcome in IPF. A previous transcriptomic prognostic biomarker has been developed that can predict outcome in IPF using blood expression data from 52 genes.

*Added value of this study:* By utilising new methods of data co-normalisation and machine learning, we were able to combine multiple publicly available datasets and perform one of the largest transcriptomic studies in IPF to-date with a total of 416 IPF cases across the discovery and validation stages. We identified three clusters of patients, one of which appeared to contain, on average, the healthiest subjects with favourable lung function and survival over time. These clusters were defined using expression from groups of genes that were significantly enriched for many different biological pathways and processes, including metabolic changes, apoptosis, cell cycle and immune response, and so could be representative of distinct pathophysiological states. Additionally, we developed a 13-gene expression-based classifier to assign individuals with IPF to one of the clusters and validated this classifier using three additional independent cohort of IPF patients (totalling 194 IPF cases). As the clusters were associated with survival, our classifier could potentially be used to predict outcome in IPF.

*Implications of all the available evidence:* Our findings support the hypothesis that the disease consists of multiple endotypes. The clusters identified in this study could provide some valuable insight into the underlying biological processes that may be driving the considerable clinical heterogeneity in IPF. With further development, our gene expression-based classifier could be a useful tool for patient stratification and outcome prediction in IPF.

## Introduction

Idiopathic pulmonary fibrosis (IPF) is a complex, ultimately fatal disease, characterised by progressive scarring of the lungs, with a median survival of 3-5 years post-diagnosis.^1,2^ Currently, there is no cure for IPF and the two drugs approved for treatment (nintedanib and pirfenidone) only slow disease progression, do not work in all patients, and are often not well tolerated.^3,4^ The clinical course of IPF is highly variable with slow progression in some patients, rapid progression in others, whilst many experience a slowly progressive course interspersed with periods of rapid lung function deterioration.^1^ It is plausible that these clinical phenotypes could reflect different disease endotypes.

Disease endotypes are subtypes of a disease as defined by a particular pathophysiological mechanism. It has been speculated that distinct endotypes of IPF exist^5,6^, as in asthma and lung cancer,^7,8^ though these are not yet well understood. Identification of endotypes would greatly increase our understanding of the behaviour and heterogeneity of the disease, and may allow for the development of biomarkers and more precise, tailored approaches to treatment.

Transcriptomic data can be used to define disease endotypes, as similar transcriptomic profiles in affected individuals may reflect common underlying biological mechanisms. Previous transcriptomic analyses of cancer patients have been particularly successful in defining clinically significant patient subgroups, which have led to improvements in treatment.^9,10^ Previous studies in IPF have used clustering of transcriptomic data to identify subtypes of patients, with smaller studies (n<200 IPF cases) generally identifying two subgroups of IPF patients and one larger study identifying three clusters of patients.

In this study, we aimed to identify clinically distinct groups of IPF patients through the application of unsupervised clustering to multiple publicly available transcriptomic datasets. We hypothesised that these groups could represent individuals with different endotypes of IPF. Rather than undertake single dataset analyses, we co-normalised and pooled multiple datasets together to increase the sample size and enhance statistical power. Additionally, we used classification to develop a method to accurately assign additional individuals with IPF to one of these groups. This classifier displayed the ability to predict survival in IPF and so we then compared the performance of our classifier in independent validation datasets to a previous method of outcome prediction in IPF.

## Methods

### Collection of publicly available data

The design of our study is shown in Figure 1. First, we reviewed the IPF datasets available on the Gene Expression Omnibus (GEO) and systematically selected several suitable datasets of gene expression data measured from whole blood (see online supplement for details).^11^ The datasets were then assigned to either the discovery stage or the validation stage (online supplement). Cohorts used in the discovery stage must have included healthy controls to enable the data co-normalisation. The methods used to pre-process the transcriptomic data before the co-normalisation are described in the online supplement.

**Figure 1:**
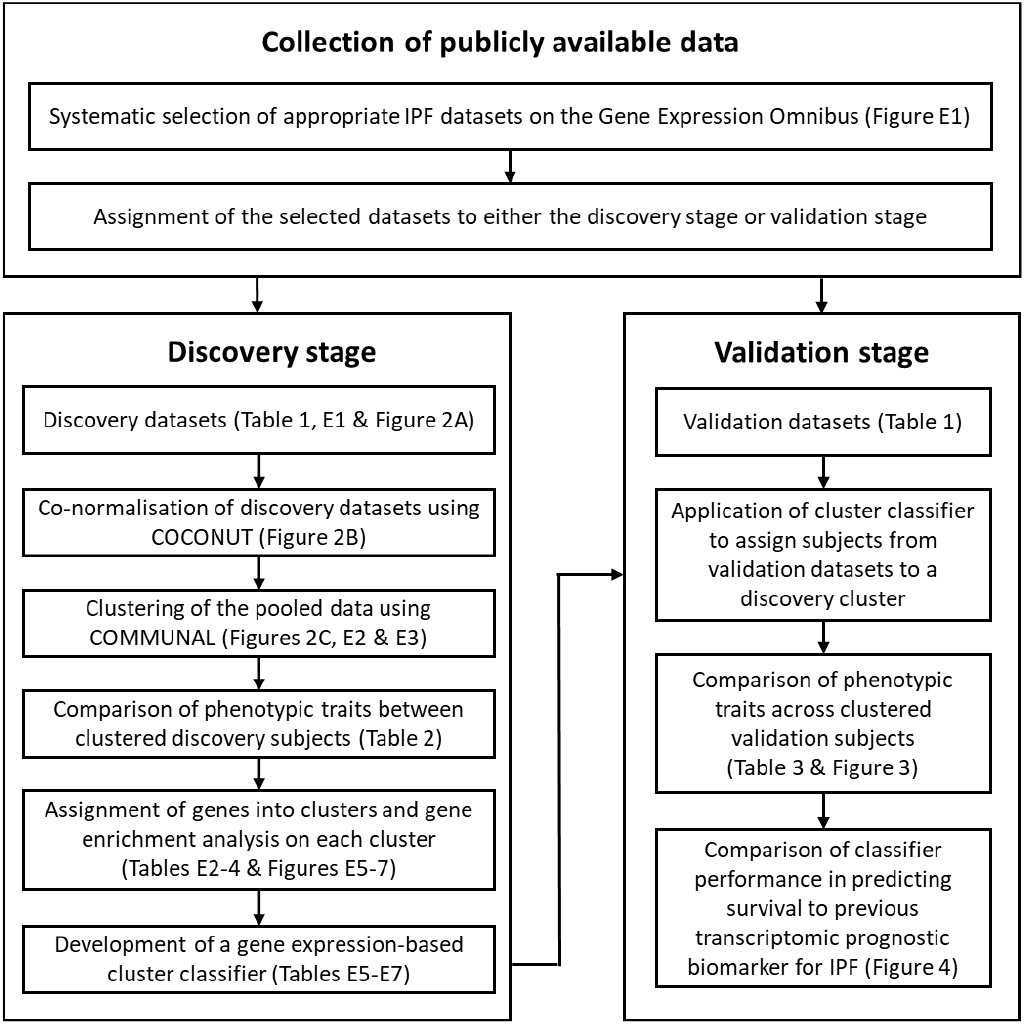
A flowchart showing the design of our study.

### Discovery stage

As the discovery datasets originated from different studies and the transcriptomic data was collected using varying platforms, there would have been considerable technical (non-biological) differences in gene expression between them. As such, the discovery datasets required adjustment before they could be combined and clustered. We co-normalised the discovery datasets using the COmbat CO-Normalization Using conTrols (COCONUT) method^12^, using R v4.0.0 and the ‘COCONUT’ package v.1.0.2 (online supplement). All healthy control subjects were then removed from further analysis.

We used the Combined Mapping of Multiple clUsteriNg ALgorithms (COMMUNAL) package v1.1.0 to identify the optimal number of clusters within the pooled, co-normalised data, using R v.3.4.0.^13^ COMMUNAL integrates data from multiple clustering algorithms across a range of genes and evaluates the validity of each number of clusters using multiple validity measures. Details on the configuration of COMMUNAL used in this study and the process used to determine the optimal cluster assignment can be found in the online supplement. Once an optimal cluster assignment was chosen, principal components analysis and heatmaps were used to visualise the separation of the clusters. Unclustered samples were excluded from further analysis.

Clinical and demographic characteristics of clustered subjects were compared using chi-square tests for count data, analysis of variance for non-skewed continuous data, Kruskal-Wallis tests for skewed continuous data and survival analysis methods for time-to-event data (online supplement). Gene enrichment analysis was performed in R v4.0.0 with the in-house ‘metabaser’ package (database v20.3, package v4.2.3) to highlight biological mechanisms that were significantly enriched for the subjects in each cluster (online supplement).

We developed a gene expression-based classifier to assign new individuals with IPF to one of the clusters using only the most informative differentially expressed genes. This classifier was designed following the approach described by Sweeney *et al*. in their study of bacterial sepsis (online supplement).^14^

### Validation stage

The classifier was used to assign all IPF subjects in each validation dataset to a discovery cluster. Phenotypic traits were compared across clusters, as in the discovery stage (online supplement).

We compared the classifier’s performance at predicting survival in IPF to a previous transcriptomic prognostic biomarker for IPF by Herazo-Maya *et al*.^15^ Each of the validation subjects with survival data available were assigned into a ‘high-risk’ or ‘low-risk’ group (in terms of mortality or requiring a lung transplant) using the method described by Herazo-Maya *et al*., the Scoring Algorithm for Molecular Subphenotypes (SAMS). For this we used as many of the genes in their signature as were present in the validation datasets. Similarly, each subject was assigned into one of our discovery clusters, which were each classed as low/high-risk based on the discovery stage findings. Survival analysis methods were used to determine which method performed best at predicting survival (online supplement).

## Results

### Collection of publicly available data

Six independent whole blood gene expression datasets were selected for inclusion in the analysis (Figure E1). Summary statistics for all subjects are shown in Table 1.

**TABLE 1:**
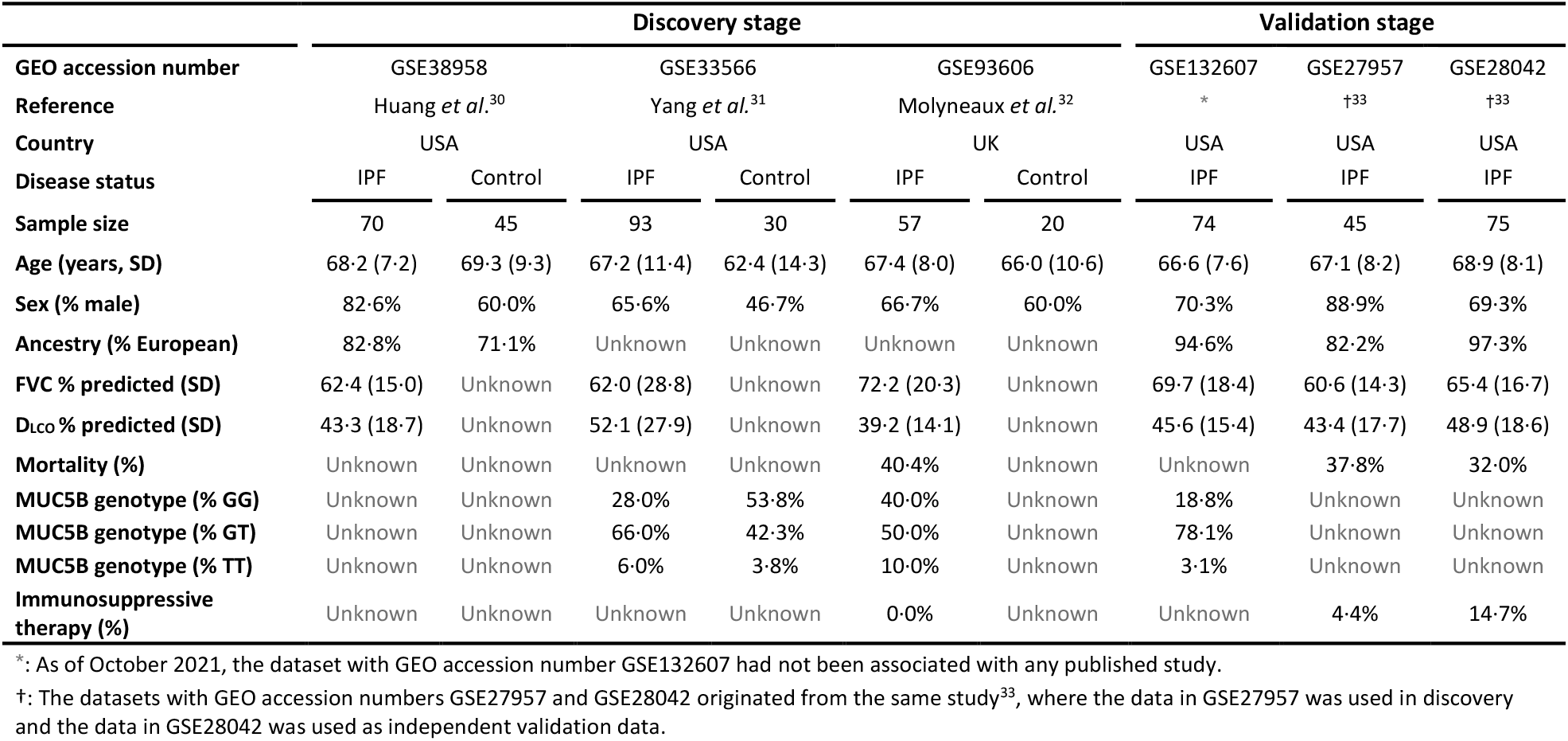
Summary information on the publicly available datasets that were included in this study, as well as summary statistics for all individuals whose data were included in the analysis. FVC = forced vital capacity, D_LCO_ = diffusing capacity of lung for carbon monoxide, SD = standard deviation, MUC5B genotype = genotype for the MUC5B promoter polymorphism rs35705950.

|  | Discovery stage |  |  |  |  |  | Validation stage |  |  |
| --- | --- | --- | --- | --- | --- | --- | --- | --- | --- |
| GEO accession number | GSE38958 |  | GSE33566 |  | GSE93606 |  | GSE132607 | GSE27957 | GSE28042 |
| Reference | Huang *et al.*<sup>30</sup> |  | Yang *et al.*<sup>31</sup> |  | Molyneaux *et al.*<sup>32</sup> |  | * | †<sup>33</sup> | †<sup>33</sup> |
| Country | USA |  | USA |  | UK |  | USA | USA | USA |
| Disease status | IPF | Control | IPF | Control | IPF | Control | IPF | IPF | IPF |
| Sample size | 70 | 45 | 93 | 30 | 57 | 20 | 74 | 45 | 75 |
| Age (years, SD) | 68.2 (7.2) | 69.3 (9.3) | 67.2 (11.4) | 62.4 (14.3) | 67.4 (8.0) | 66.0 (10.6) | 66.6 (7.6) | 67.1 (8.2) | 68.9 (8.1) |
| Sex (% male) | 82.6% | 60.0% | 65.6% | 46.7% | 66.7% | 60.0% | 70.3% | 88.9% | 69.3% |
| Ancestry (% European) | 82.8% | 71.1% | Unknown | Unknown | Unknown | Unknown | 94.6% | 82.2% | 97.3% |
| FVC % predicted (SD) | 62.4 (15.0) | Unknown | 62.0 (28.8) | Unknown | 72.2 (20.3) | Unknown | 69.7 (18.4) | 60.6 (14.3) | 65.4 (16.7) |
| D<sub>LCO</sub> % predicted (SD) | 43.3 (18.7) | Unknown | 52.1 (27.9) | Unknown | 39.2 (14.1) | Unknown | 45.6 (15.4) | 43.4 (17.7) | 48.9 (18.6) |
| Mortality (%) | Unknown | Unknown | Unknown | Unknown | 40.4% | Unknown | Unknown | 37.8% | 32.0% |
| MUC5B genotype (% GG) | Unknown | Unknown | 28.0% | 53.8% | 40.0% | Unknown | 18.8% | Unknown | Unknown |
| MUC5B genotype (% GT) | Unknown | Unknown | 66.0% | 42.3% | 50.0% | Unknown | 78.1% | Unknown | Unknown |
| MUC5B genotype (% TT) | Unknown | Unknown | 6.0% | 3.8% | 10.0% | Unknown | 3.1% | Unknown | Unknown |
| Immunosuppressive therapy (%) | Unknown | Unknown | Unknown | Unknown | 0.0% | Unknown | Unknown | 4.4% | 14.7% |

\*: As of October 2021, the dataset with GEO accession number GSE132607 had not been associated with any published study.
†: The datasets with GEO accession numbers GSE27957 and GSE28042 originated from the same study<sup>33</sup>, where the data in GSE27957 was used in discovery and the data in GSE28042 was used as independent validation data.

### Discovery stage

All three discovery stage datasets were microarray-based (Table E1). There were expression levels measured for 9,371 common genes across the three datasets, which consisted of a total of 220 IPF subjects and 95 healthy control subjects.

Prior to COCONUT co-normalisation, the data from the three cohorts were entirely separated in high-dimensional space due to technical differences between the studies (Figure 2A). Whereas after COCONUT (Figure 2B), the data was overlapping in high-dimensional space, indicating that the technical differences between datasets had been reduced and that the co-normalised data was suitable for clustering.

**Figure 2:**
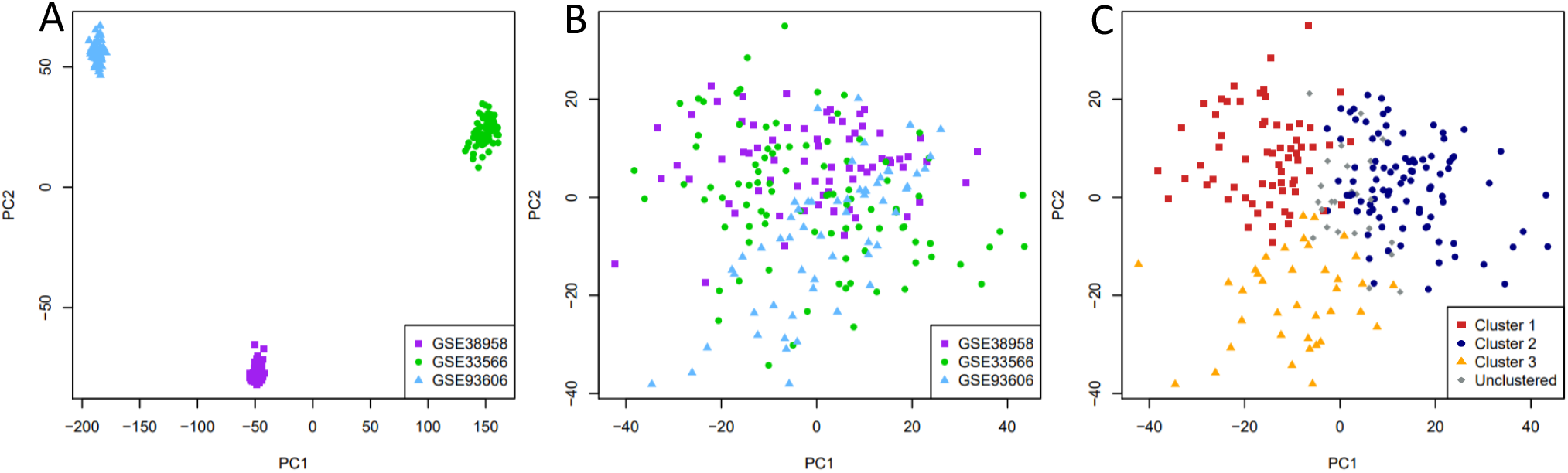
Plots of the first two principal components of the gene expression data for the IPF samples prior to co-normalisation and stratified by original study (A), post co-normalisation and stratified by original study (B) and post co-normalisation stratified by cluster (C). The x-axis represents the first principal component of the data and the y-axis represents the second principal component of the data.

COMMUNAL was run on the co-normalised data and the resulting optimality map is shown in Figure E2. The clustering assignment with 3 clusters using 2,500 genes was chosen as the optimal assignment (online supplement), with 64 subjects assigned to Cluster 1, 95 assigned to Cluster 2, 37 assigned to Cluster 3 and 24 (10·4%) that were unclustered (Figure 2C and Figure E3).

Table 2 shows the clinical and demographic traits of the subjects in each cluster by study, as well as for all studies combined. With all studies combined, statistically significant differences in average predicted Diffusing capacity of the Lung for carbon monoxide (DL_CO_) were observed across clusters (P=0·009). Subjects in Cluster 1 had a similar median predicted DL_CO_ to those in Cluster 3, whilst subjects in Cluster 2 had the greatest median predicted DL_CO_, indicating that these individuals had relatively preserved lung function. Additionally, there was a significant difference in average score from the Gender, Age and Physiology (GAP) index for IPF mortality (P=0·006),^16^ with those in Cluster 1 having the greatest GAP score and those in Cluster 2 having the lowest average GAP score. There was a statistically significant difference in mortality between Clusters 2 and 3, with death observed for 25% of subjects in Cluster 2 and 67% of subjects in Cluster 3 (P=0·009). Furthermore, those in Cluster 3 had consistently poorer survival over time than those in Cluster 2 (Figure E4). A Cox proportional-hazards (PH) model estimated that the hazard ratio (HR) between Clusters 2 and 3 was 3.59 (95% CI: [1·40, 9·19], P=0·008), and so at any follow-up time, subjects in Cluster 3 were estimated to be 3·59 times as likely to die as subjects in Cluster 2.

**TABLE 2:** Comparison of clinical and demographic traits of clustered subjects by study and for all studies combined. Data are presented as count (percentage), mean (standard deviation, SD) or median (interquartile range, IQR). NA = data not available, FVC=Forced vital capacity, D_LCO_ = Diffusing capacity for carbon monoxide, FEV_1_ = Forced expiratory volume in one second, CPI = composite physiologic index, MUC5B genotype = genotype for the MUC5B promoter polymorphism rs35705950. - indicates that the calculation was not applicable as there were zero subjects in that cluster. P-value for count data is from a chi-square test, test comparing means is analysis of variance and test comparing medians is the Kruskal-Wallis log rank test. Significant P-values (P < 0·05) are highlighted in bold.

|  | GSE38958 (n=65) |  |  | GSE33566 (n=83) |  |  | GSE93606 (n=48) |  |  | All studies combined (n=196) |  |  |  |  |
| --- | --- | --- | --- | --- | --- | --- | --- | --- | --- | --- | --- | --- | --- | --- |
|  | Cluster 1 | Cluster 2 | Cluster 3 | Cluster 1 | Cluster 2 | Cluster 3 | Cluster 1 | Cluster 2 | Cluster 3 | Cluster 1 | Cluster 2 | Cluster 3 | P-value | Total n used |
| n subjects in cluster | 22 | 39 | 4 | 42 | 32 | 9 | 0 | 24 | 24 | 64 | 95 | 37 |  |  |
| Age (years)<br>(mean, SD) | 70.0<br>(6.3) | 68.3<br>(7.9) | 64.0<br>(2.7) | 66.7<br>(9.8) | 67.0<br>(14.1) | 67.0<br>(12.1) | - | 64.8<br>(5.9) | 70.3<br>(8.8) | 67.8<br>(8.9) | 66.9<br>(10.2) | 68.8<br>(9.4) | 0.592 | 188 |
| Male<br>(%) | 20<br>(91.0%) | 30<br>(77.0%) | 4<br>(100%) | 32<br>(76.2%) | 21<br>(65.6%) | 3<br>(33.3%) | - | 15<br>(62.5%) | 16<br>(66.7%) | 52<br>(81.3%) | 66<br>(69.5%) | 23<br>(62.2%) | 0.091 | 196 |
| European ancestry<br>(%) | 17<br>(81.0%) | 29<br>(82.9%) | 3<br>(75.0%) | NA | NA | NA | - | NA | NA | 17<br>(81.0%) | 29<br>(82.9%) | 3<br>(75.0%) | 0.883 | 60 |
| Ever smoker<br>(%) | NA | NA | NA | NA | NA | NA | - | 15<br>(62.5%) | 18<br>(78.3%) | NA | 15<br>(62.5%) | 18<br>(78.3%) | 0.389 | 47 |
| Death observed<br>during study (%) | NA | NA | NA | NA | NA | NA | - | 6<br>(25.0%) | 16<br>(66.7%) | NA | 6<br>(25.0%) | 16<br>(66.7%) | **0.009** | 48 |
| FVC % predicted<br>(median, IQR) | 59.5<br>(19.5) | 65.0<br>(24.0) | 51.5<br>(7.8) | 77.0<br>(36.0) | 66.0<br>(46.0) | 73.0<br>(17.5) | - | 71.5<br>(27.7) | 60.8<br>(24.1) | 63.0<br>(35.0) | 70.5<br>(30.1) | 60.1<br>(23.4) | 0.342 | 154 |
| D<sub>LCO</sub> % predicted<br>(median, IQR) | 34.5<br>(17.5) | 49.0<br>(21.0) | 28.5<br>(21.0) | 65.0<br>(37.0) | 66.0<br>(40.0) | 30.0<br>(30.0) | - | 38.1<br>(17.1) | 36.6<br>(15.9) | 35.0<br>(30.0) | 45.0<br>(29.2) | 34.4<br>(17.3) | **0.009** | 133 |
| FEV<sub>1</sub> % predicted<br>(median, IQR) | NA | NA | NA | NA | NA | NA | - | 74.9<br>(23.1) | 65.4<br>(22.7) | NA | 74.9<br>(23.1) | 65.4<br>(22.7) | 0.216 | 48 |
| GAP index<br>(mean, SD) | 5.3<br>(1.3) | 3.9<br>(1.3) | 4.5<br>(1.3) | 4.3<br>(1.5) | 4.1<br>(1.6) | 4.3<br>(3.1) | - | 3.7<br>(1.8) | 4.4<br>(1.6) | 4.9<br>(1.4) | 3.9<br>(1.5) | 4.4<br>(1.7) | **0.006** | 132 |
| MUC5B genotype:<br>GG (%) | NA | NA | NA | 5<br>(29.4%) | 6<br>(28.6%) | 3<br>(60.0%) | - | 5<br>(26.3%) | 11<br>(50.0%) | 5<br>(29.4%) | 11<br>(27.5%) | 14<br>(51.9%) | 0.230 | 84 |
| MUC5B genotype:<br>GT (%) | NA | NA | NA | 10<br>(58.8%) | 14<br>(66.7%) | 2<br>(40.0%) | - | 12<br>(63.2%) | 8<br>(36.4%) | 10<br>(58.8%) | 26<br>(65.0%) | 10<br>(37.0%) |  |  |
| MUC5B genotype:<br>TT (%) | NA | NA | NA | 2<br>(11.8%) | 1<br>(4.8%) | 0<br>(0%) | - | 2<br>(10.5%) | 3<br>(13.6%) | 2<br>(11.8%) | 3<br>(7.5%) | 3<br>(11.1%) |  |  |

Gene enrichment analysis revealed that Cluster 1 was significantly enriched for biological mechanisms relating to metabolic changes (Table E2 and Figure E5). Cluster 2 was significantly enriched for biological processes and pathways relating to gene regulation, DNA repair, cell cycle and apoptosis (Table E3 and Figure E6), whilst Cluster 3 was significantly enriched for terms relating to the immune response (Table E4 and Figure E7). In addition, the genes assigned to Clusters 2 and 3 were each found to be statistically over-connected (in terms of direct gene regulation) to a significant number of genes that have been previously implicated in the development of IPF (online supplement).

We used the pooled, co-normalised gene expression data for all 196 subjects who were successfully clustered in the discovery analysis to train a gene expression-based cluster classifier (online supplement). The classifier used expression data from 13 genes (Table E5) and was able to accurately reassign 99·0% of discovery subjects (Table E7).

### Validation stage

There were 194 IPF subjects across the three validation cohorts. Expression levels for all 13 genes used in the classifier were available in all three validation cohorts. We used the classifier to assign each individual to a cluster and compared phenotypic traits across clusters (Table 3). As in the discovery stage, there were statistically significant differences in mortality between clusters (P=0·001) and those in Cluster 2 had the best survival over time (Figure 3). Additionally, individuals in Cluster 2 had the highest average DL_CO_, though the difference in DL_CO_ between validation clusters was not statistically significant (P=0·069). Cox PH models (Table E8) estimated that at any follow-up time, an individual in Cluster 1 was 3.80 times more likely to die than an individual in Cluster 2 (95% CI = [1·78, 8·12], P=0·001), whilst an individual in Cluster 3 was 5·05 times more likely to die than an individual in Cluster 2 (95% CI = [2·24, 11·35], P=9·1×10^−5^). However, the difference in survival over time between Clusters 1 and 3 was not statistically significant (HR= 1·47 (95% CI [0·67, 3·22], P=0·341).

**TABLE 3:**
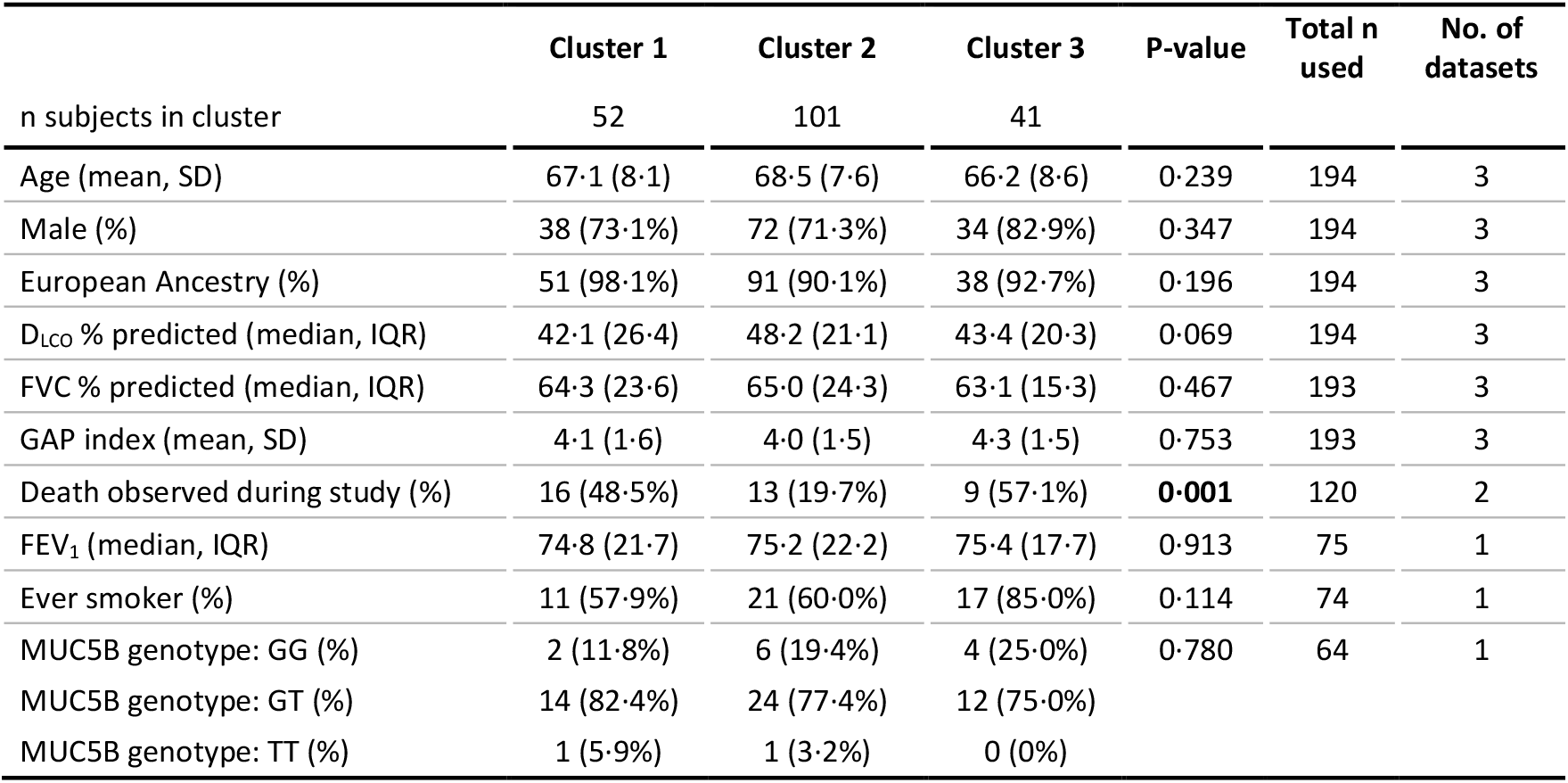
Comparison of phenotypic traits across clusters when all validation subjects were clustered using the cluster classifier. Data are presented as count (percentage), mean (standard deviation, SD) or median (interquartile range, IQR). FVC=Forced vital capacity, D_LCO_ = Diffusing capacity for carbon monoxide, CPI = composite physiologic index, FEV_1_ = Forced expiratory volume in one second, MUC5B genotype = genotype for the MUC5B promoter polymorphism rs35705950. Significant P-values (P < 0·05) are highlighted in bold.

**Figure 3:**
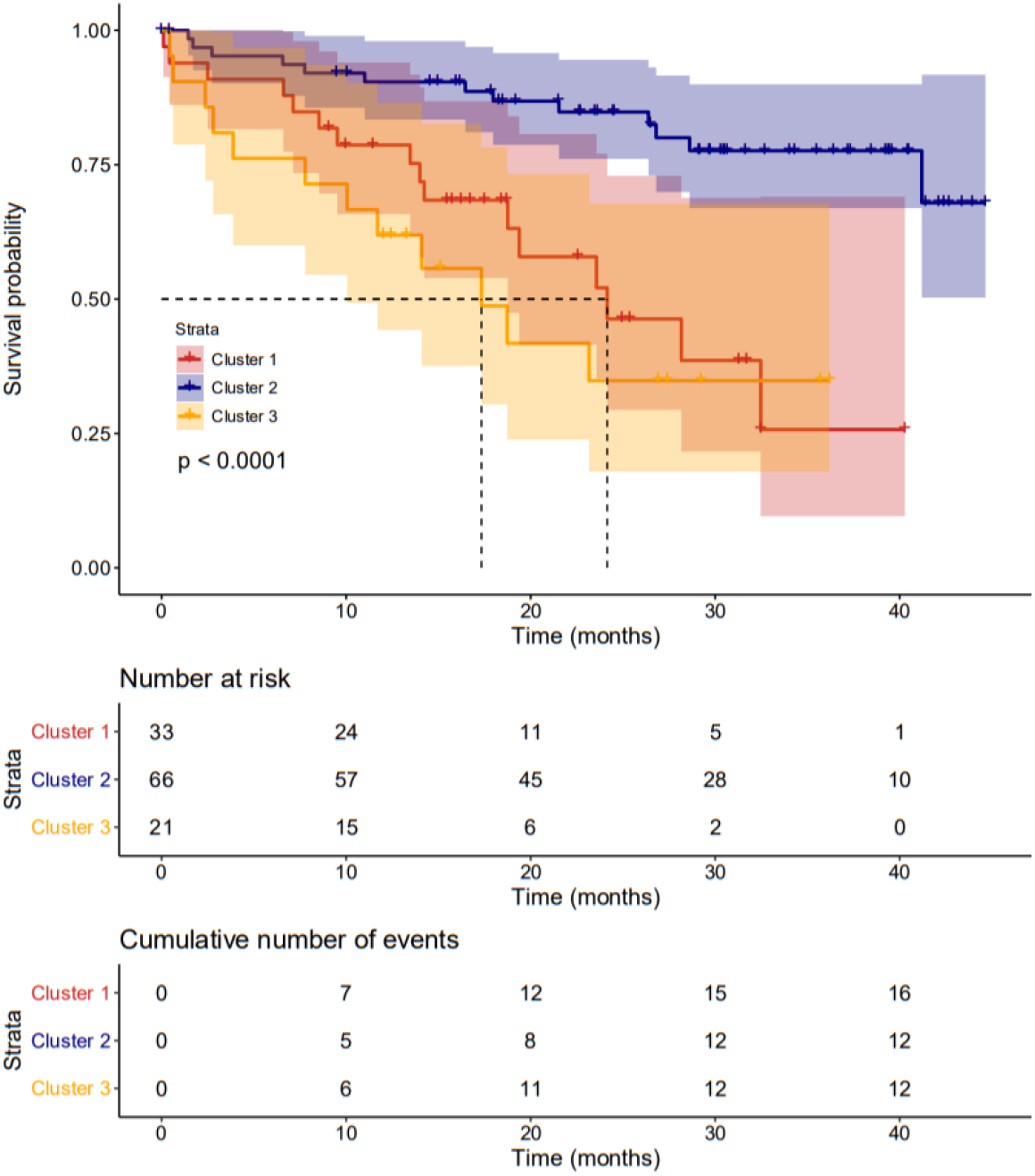
A Kaplan-Meier plot showing survival over time for the clustered validation subjects. The p-value shown on the plot is from a log-rank test testing the three curves for equality. Median survival in each cluster is shown by dotted lines, where possible.

Finally, we compared the performance of our classifier at predicting survival in IPF to SAMS, a method used by Herazo-Maya *et al*. to predict outcome in IPF using a 52-gene signature.^15^ There were no common genes between the classifier and the 52-gene signature, though many were highly correlated in the validation subjects (Figure E8). The subjects in the GSE27957 and GSE28042 validation cohorts (GSE132607 did not report mortality) were each classed as ‘high-risk’ or ‘low-risk’ using both gene expression-based methods. As Clusters 1 and 3 were not significantly distinct in terms of survival, both clusters were considered equally ‘high risk’ for the assignments based on the 13-gene classifier. 51/52 (98·1%) genes in Herazo-Maya *et al*.’s gene signature were present in the GSE27957 dataset and 50/52 (96·2%) were available in the GSE28042 dataset. Overall, there was 68·3% agreement between the two methods (Table E9).

Our classifier performed well at predicting survival (Figure 4A), with the subjects in the ‘high-risk’ clusters having far poorer survival over time than those in the ‘low-risk’ cluster. A univariate Cox PH model estimated that at any follow-up time, an individual in a high-risk cluster was 4·25 times more likely to die than an individual in the low-risk cluster (95% CI = [2·14, 8·46], P=3·7×10^−5^). This model had a C-index (the equivalent of the area under the curve [AUC] for a receiver operating characteristic [ROC] curve) of 0·664 (95% CI= [0·590, 0·737]). SAMS (Figure 4B) performed less well, with a Cox PH model estimating that at any time, those in the high-risk group were 1.98 times as likely to die than those in the low-risk group (95% CI = [1·07, 3·68], P = 0·030) and a C-index of 0·609 (95% CI = [0·531, 0·686]).

**Figure 4:**
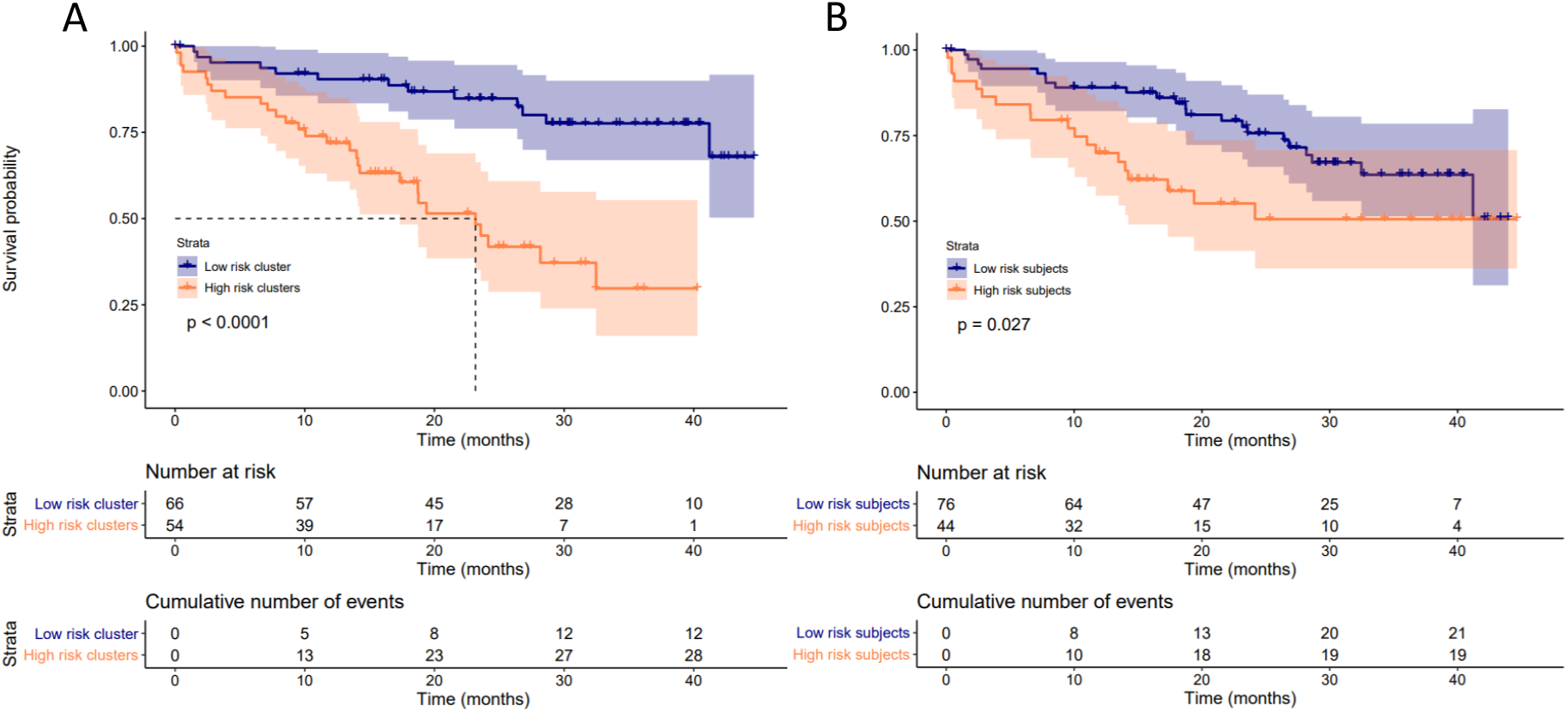
Survival over time for the IPF subjects in GSE27957 and GSE28042, stratified by risk group according to our 13 gene classifier (A) and Herazo-Maya *et al*.’s method SAMS (B). The P-value on each plot is from a log-rank test testing the two curves for equality. A dotted line on the plot indicates the median survival time for the risk group if this could be calculated.

The risk predictions made using the classifier remained statistically significant (P=0·007) after adjusting for age, sex, ancestry, FVC and DL_CO_ (Table E10), with a hazard ratio of 2·70 between the high-risk and low-risk clusters (95% CI= [1·32, 5·53]). This model had a C-index of 0·773 (95% CI = [0·697, 0·848]), which was greater than that of the Cox model containing only age, sex, ancestry, FVC and DL_CO_ (C-index = 0·747, 95% CI = [0·670, 0·825]), suggesting an improvement in predictive ability. A likelihood ratio test between the two models gave a P-value of 0·005, suggesting that the improvement in predictive ability when including the classifier’s risk predictions was statistically significant. The multivariate Cox model containing SAMS’ risk predictions had a C-index of 0·760 (95% CI = [0·684, 0·837]), which suggested an improvement over the Cox model containing only age, sex, ancestry, FVC and DL_CO_, though the likelihood ratio test p-value between these two models was not statistically significant (P=0·105).

## Discussion

By applying new statistical methods for data co-normalisation and machine learning to multiple publicly available datasets, we identified three clusters of IPF patients with statistically significant differences in lung function and survival. As the clustering in this study was undertaken independently of clinical data, yet significant differences in clinical traits were observed between clusters, this suggests that they may be representative of distinct and clinically relevant endotypes of IPF.

In this study we used datasets in which the gene expression had been measured from whole blood samples. However, as IPF is a lung disease, characterised by damage to the alveolar epithelium, patterns of gene expression identified in blood may not reflect the underlying pathology of the disease and may instead reflect downstream effects or the presence of confounders, such as secondary infections or treatment effects. Nonetheless, blood is more accessible than a lung-specific tissue/cell type and the expression of a gene in blood is often a significant predictor of the expression of that gene in lung.^17^ Furthermore, the blood expression datasets available on GEO provided a larger sample size and more comprehensive accompanying clinical data than lung-specific tissue types, which allowed us to identify statistically significant clinical differences between clusters. In addition, this allowed us to develop a blood-based classifier, which has more clinical utility than one that requires measurements from lung, as this would require more invasive sample collection.

The genes that were most differently expressed in subjects in Cluster 1 were significantly enriched for biological mechanisms related to metabolic changes. Recent findings appear to suggest that metabolic dysregulation could be a contributing factor to fibrosis, though its role is not yet fully understood.^18,19^ The genes in Cluster 1 were also significantly enriched for pathways related to TGF-β signalling, which is a central mediator of fibrosis.^20–22^

Among the biological pathways that were significantly enriched for Cluster 2 were pathways related to apoptosis and cell cycle. It has been previously reported that apoptosis is increased in alveolar epithelial cells of IPF patients but decreased in myofibroblasts,^23^ with this imbalance contributing to IPF pathogenesis.^24^ Furthermore, the use of therapies that can selectively manipulate apoptosis have been proposed.^25^ Additionally, genetic variants within cell cycle genes have been shown to be associated with IPF development and progression.^26^ The results for this cluster could further support the idea that apoptosis and cell cycle each play an important role in the pathology of IPF.

Cluster 3 was enriched for terms related to the immune system response. The role of the immune system in IPF has been controversial in the past; failed immunomodulatory therapies in IPF, some of which have led to worse outcomes, have led to speculation that certain immune responses are protective while others are harmful.^27,28^ An improved understanding of immune-driven endotypes could inform novel treatment approaches.

The 13-gene expression-based cluster classifier was successfully validated as it was able to assign the IPF subjects from the validation datasets to clusters with statistically significant differences in survival between groups that were consistent with the discovery clusters. As the classifier had the ability to assign subjects who are at a lower risk of death into Cluster 2 and the subjects who are at a greater risk of death into the other two clusters, it could potentially be used to predict survival in IPF.

The performance of the classifier in predicting survival was compared with SAMS, a similar approach to outcome prediction in IPF.^15^ Despite using data from one quarter of the number of genes used for SAMS, the differences in survival over time observed between the risk groups in the two validation datasets had greater statistical significance and effect size when predictions were made using the classifier. Additionally, including the classifier’s predictions in a survival model that adjusted for important covariate factors led to a statistically significant increase in predictive ability.

One of the main strengths of this study was that the utilization of a new statistical approach to co-normalisation (COCONUT) allowed for three datasets to be combined,^12^ resulting in one of the largest transcriptomic studies in IPF to date with a total of 414 IPF cases across the discovery and validation stages. Another strength of our study was that the application of COMMUNAL, which considered two different clustering algorithms and tested five validity measures over a range of genes, meant that our clustering was more reliable and more likely to be reproducible than the standard approach, which would have been to apply one clustering algorithm and test one validity measure.

There were several limitations to this study. Firstly, as we relied on the use of publicly available data, some clinical variables were relatively underpowered due to missingness within the data or having not been reported in all studies. In addition, we lacked detailed data for clinically significant traits such as patient reported outcomes, lung function decline over time and did not possess information regarding the background therapy of the IPF subjects. However, for the three cohorts with survival data available, we were able to glean from the original papers that the IPF patients were either treatment naïve populations (GSE93606) or that there were only a small proportion that were receiving immunosuppressive therapy at the time of the blood collection (GSE27957 and GSE28042). In addition, these populations were not given anti-fibrotics and so treatment effects are unlikely to have been driving the large differences in survival that were observed between clusters.

A further weakness of our study is that each participating cohort of IPF subjects was subject to survival bias, as only subjects who survived long enough to enrol into each study could have contributed their transcriptomic data to it. This could have restricted the level of heterogeneity of IPF that we were able to capture in the study and limited the generalisability of our findings.

Additionally, COCONUT makes the assumption that the healthy controls across the different studies came from the same statistical distribution and so all differences between healthy controls across studies must have been due to non-biological variation. This means that any large differences in confounding factors between the groups of healthy controls would have restricted the efficacy of the co-normalisation.

If the clusters identified in this study do truly represent endotypes of IPF, it may be worth speculating about the nature of these endotypes. As IPF is a complex disease, with many known common genetic and environmental exposures, it is unlikely that it would behave under a traditional discrete endotype model and instead more likely that it would behave under a more complex model, such as the palette model described by McCarthy.^29^ Our gene enrichment analysis results could implicate metabolic changes and the immune system response as being among the component pathways for IPF.

To conclude, these results could support the hypothesis of multiple endotypes of IPF as there appear to be at least two clinically distinct groups of IPF patients that can be identified through cluster analysis of transcriptomic data. As these clusters were defined using expression from groups of genes that were significantly enriched for many different biological pathways and processes, they could be representative of distinct pathophysiological states. Additionally, a classifier with the ability to assign additional individuals with IPF to one of the clusters was developed. With further development, this classifier could be a useful tool in outcome prediction in IPF as well as helping us gain a better understanding of the underlying biological processes that may be driving the observed differences in survival.

## Data Availability

The data that supports the findings in this study are openly available via the Gene Expression Omnibus (https://www.ncbi.nlm.nih.gov/geo/).

https://www.ncbi.nlm.nih.gov/geo/query/acc.cgi?acc=GSE38958

https://www.ncbi.nlm.nih.gov/geo/query/acc.cgi?acc=GSE33566

https://www.ncbi.nlm.nih.gov/geo/query/acc.cgi?acc=GSE93606

https://www.ncbi.nlm.nih.gov/geo/query/acc.cgi?acc=GSE132607

https://www.ncbi.nlm.nih.gov/geo/query/acc.cgi?acc=GSE27957

https://www.ncbi.nlm.nih.gov/geo/query/acc.cgi?acc=GSE28042

## Acknowledgements

We thank the research teams who have made their data publicly available via the Gene Expression Omnibus and to all study participants for contributing their data and samples.

