## Supplementary material for "Cluster analysis of transcriptomic datasets to identify endotypes of Idiopathic Pulmonary Fibrosis": Online supplement

### Cluster analysis of transcriptomic datasets to identify endotypes of Idiopathic Pulmonary Fibrosis – online data supplement

Luke M. Kraven[1,2*], Adam R. Taylor[2*], Philip L. Molyneaux[3,4], Toby M. Maher[3,4,5], John E. McDonough[6], Marco Mura[7], Ivana V. Yang[8], David A. Shwartz[8], Yong Huang[9], Imre Noth[9], Shwu-Fan Ma[9], Astrid J. Yeo[2*], William A. Fahy[2*], R. Gisli Jenkins[3,4*], Louise V. Wain[1,10*]

#### Additional text

##### Systematic selection of publicly available datasets

We performed our systematic search in March 2020 to select the datasets that were suitable for inclusion in the study (Figure E1). We required multiple sets of transcriptomic data from independent cohorts. We searched the Gene Expression Omnibus (GEO) (1) for all collections that contained the term 'IPF', excluding any that did not contain human samples. We restricted the search to collections with at least 30 samples as this allowed for inclusion of the largest datasets with the most IPF cases and healthy control subjects, which are the datasets that were the most likely to successfully co-normalise due to the higher counts of healthy control subjects. We did not restrict the search by platform. Each of the remaining collections were then reviewed to assess whether they contained data for IPF cases. All collections that did not contain data for IPF subjects were excluded.

For a successful co-normalisation and meaningful clustering results, we were required to choose an optimal tissue/cell type to use for the analysis. After reviewing the IPF datasets on GEO, we chose whole blood as our optimal tissue/cell type. There were three main reasons for this. Firstly, there were several relatively large whole blood datasets available on GEO and these would have provided the largest sample size and greatest statistical power for the study compared to other tissue types. Secondly, we required multiple datasets that contained data for healthy controls in addition to the IPF patients (so that the data could be co-normalised using COCONUT) and the whole blood datasets fulfilled this requirement. Thirdly, the accompanying clinical data for the whole blood datasets was far more comprehensive than for other tissue types, such as whole lung. This clinical data was vital to the study as it was required for the characterisation of the clusters in both the discovery and validation stages. So, all GEO collections containing expression data measured from a non-blood tissue/cell type were excluded.

As multiple transcriptomic datasets were to be combined, it was important to check for the presence of common individuals across cohorts, which would have meant that the cohorts were not independent and could have biased the results of the study. To this end, the subjects in each collection were checked for unique study identification codes. Using these, we found that two of the blood collections, GSE132607 (n=74) and GSE85268 (n=68), both contained subjects from the Correlating Outcomes With Biochemical Markers to Estimate Time-progression in Idiopathic Pulmonary Fibrosis (COMET) study (ClinicalTrials.gov identifier: NCT01071707). There were a large number of IPF subjects in common between the two cohorts (n=58) and so we excluded the GSE85268 dataset as it was the collection with fewer IPF subjects.

The seven remaining collections of data were uploaded by research groups from across the USA (including the University of Virginia, Yale University, the University of Nevada and the University of Colorado) and the UK (Imperial College London). GSE27957 and GSE28042 were uploaded by the Kaminski Lab in Yale. These two collections were both used in the same study (2), where GSE27957 was used as discovery data and GSE28042 was used as independent replication data. Similarly, the data found in GSE133298 and GSE132607 were uploaded by researchers at the University of Virginia and were used as independent cohorts in the same study (unpublished as of October 2020, both collections uploaded to GEO in September 2019). All remaining collections were uploaded by separate research groups and no additional evidence of common subjects across cohorts was found so the seven cohorts of IPF subjects were deemed independent. However, the possibility that subjects could be common in two or more studies cannot be ruled out.

The human biological samples were sourced ethically and their research use was in accord with the terms of the informed consents under an institutional review board/ethical committee (IRB/EC)-approved protocol.

##### Assignment of datasets to discovery and validation stages

All cohorts included in the discovery stage must have contained healthy controls in order to enable the data co-normalization step. Four of the seven selected blood datasets contained data for healthy controls. We used the three with the greatest number of controls in discovery as these were the most likely to successfully co-normalize. The four remaining datasets were reserved for use in the validation stage. One dataset (GSE133298) was excluded during the validation stage as not all of the genes that were required to fully apply the classifier were present in the dataset.

##### Data pre-processing

In each discovery dataset, probes that did not map to a gene were removed. In the instance where multiple probes mapped to the same gene, only the probe with the greatest mean expression was included in the analysis. Each dataset was then quantile normalised to reduce any technical differences between the gene probes within a study. Following this, each dataset was scaled so that all expression data was on the $\log_2$ scale and thus in a consistent form prior to co-normalisation. Genes were matched across studies based on their gene symbols.

##### Data co-normalisation using COCONUT

We used COmbat CO-Normalization Using conTrols (COCONUT) (3) (in R v4.0.0 and the 'COCONUT' package) to reduce the technical differences between the three discovery transcriptomic datasets, therefore enabling a cluster analysis to be performed on the pooled, co-normalized data. COCONUT is an unbiased co-normalisation method which assumes that all healthy controls across studies come from the same statistical distribution. It uses the healthy controls in each study to calculate correction factors that remove the technical differences in the data for the diseased subjects, without bias to the number of disease cases present. The method is adapted from the ComBat empiric Bayes normalization method (4), which is often used to adjust for batch effects within a study.

Data for each study was input into COCONUT by providing a gene expression matrix (on the $\log_2$ scale) of common genes against subjects. These were accompanied by an indicator variable that showed which subjects were cases and which were controls. Following the co-normalisation, we removed all healthy control subjects from further analysis. Plots of the first two principal components of the transcriptomic data before and after COCONUT were used to evaluate the efficacy of the co-normalisation.

##### Clustering using COMMUNAL

In this study, we ran COMMUNAL using consensus clustering versions of two algorithms, K-means clustering and partitioning around medoids (PAM). Five different metrics were used to assess the validity of the clustering for different numbers of clusters and genes. These were: the gap statistic, connectivity, average silhouette width, the G3 metric, and Pearson's gamma coefficient. We ranked the genes in order of variance, with the 'top' 100 genes referring to the 100 genes with the greatest variance. We then applied the COMMUNAL algorithm using a range of input genes from the top 100 to the top 5,000. The genes with the greatest variance were used as these were the most likely to be informative, so as to minimise the number of non-informative genes and increase the signal-to-noise ratio.

The samples that were not assigned into the same cluster by the COMMUNAL clustering algorithms were labelled 'unclustered'. Since the intention was to use the clustered data to create a classifier and classifiers trained on data with fewer errors are more robust, these uncertain samples were removed from further analysis to improve the accuracy of the classifier.

The results were visualised in the form of a 3-dimensional (3D) map (Figure E2), which we used to select the optimal number of clusters in the data, as well as the optimal number of genes to use in the clustering. The map shows the mean of standardized values of each validity measure across the entire tested space. On the 3D map, blue squares indicate a potentially optimal clustering at a certain number of genes by finding the assignment where the mean combined validation metric is greatest. The absolute maximum number of clusters for any consensus subset is marked with a red square. The points where the blue and red squares overlap indicate stable optima. If stable optima at a particular number of clusters are observed over most of the tested space, this indicates the presence of a strong, consistent biological signal at this number of clusters.

In Figure E2 there are stable optima at K=4 from 250 genes to 1,000 genes, and at K=3 from 2,500 genes to 5,000 genes, as shown by the red and blue squares meeting. Despite the K=4 clustering assignment at 1,000 genes showing the highest mean standardized validity score of all tested clustering assignments, there were stable optima at K=3 clusters over a larger range of tested space, indicating a stronger biological signal. As such, K=3 was chosen as the optimal number of clusters in the pooled IPF dataset. The clustering at 2,500 genes and 3 clusters was chosen as the optimal clustering assignment, under the assumption that the assignment with the fewest number of genes (out of those with stable optima at K=3) has the least amount of redundant signal.

##### Comparison of phenotypic traits across clusters

We characterised the clusters by comparing the clinical and demographic traits of the subjects that were assigned to each cluster. This was done for each phenotypic trait that was reported in at least one discovery cohort and one validation cohort. The statistical significance of the phenotypic differences across clusters was evaluated for all studies combined using a chi-square test for count data, an analysis of variance to compare means for non-skewed continuous data and a Kruskal-Wallis rank sum test to compare medians for skewed continuous data. For traits in the form of time-to-event data, Kaplan-Meier plots were used to approximate and visualise the survival function for these variables. Further, Cox proportional-hazards (PH) models were fit with cluster as the sole independent variable and the time to the event as the response variable.

##### Gene enrichment analysis

First, we assigned each of the 2,500 genes used in the optimal COMMUNAL clustering assignment to the cluster in which its expression was most different to its expression in the other two clusters, as this suggests that that gene was contributing to the identity of that cluster. 814 genes were assigned to Cluster 1, 866 were assigned to Cluster 2 and 820 were assigned to Cluster 3.

We then performed multiple ANOVA tests (one for each cluster) for each gene, each comparing the expression of that gene in subjects within a given cluster against the expression of subjects in both other clusters. Each gene was then assigned to the cluster in which it had the lowest ANOVA p-value. One benefit of this approach is that the ANOVA tests allowed for filtering based on statistical significance; a nominal p-value significance threshold of 0.05 was introduced and genes whose lowest ANOVA p-value was greater than this threshold were removed. The rationale for the introduction of this filtering step was that removing genes that were not associated with any cluster would reduce noise and strengthen the gene enrichment analysis for each cluster. The threshold for statistical significance was kept at a nominal level as a correction for all 7,500 ANOVA tests would have likely left too few genes assigned to each cluster to successfully perform the enrichment analysis. After the removal of the genes that were not at least nominally associated to any cluster, there were 769 genes assigned to Cluster 1, 839 assigned to Cluster 2 and 784 assigned to Cluster 3.

Then, gene enrichment analysis was performed separately on the three resulting gene lists using R v.4.0.0 and the in-house package 'metabaser' (database v20.3, package v4.2.3). This was used to search databases of gene ontology terms for statistically overrepresented *biological processes* and *biological pathways*. At the time that the analysis was performed, there were 17,552 *biological processes* and 12,222 *biological pathways* in the database accessed by metabaser. metabaser reports 'q-values', which are p-values that have been adjusted for multiple tests using the false-discovery rate. Gene ontology terms with q-value < 0.05 were deemed statistically significant. Sankey plots were used to show which of the genes that were assigned to each cluster corresponded to the 20 most significantly enriched *biological pathways* (see Figure 3).

Additionally, the gene lists of each cluster were searched for the presence of the nearest gene for any of the 14 variants that were genome-wide significant in Allen et al. (5), the largest genome-wide association study meta-analysis of IPF susceptibility to-date. The 14 genes were as follows: *AKAP13*, *ATP11A*, *DEPTOR*, *DPP9*, *DSP*, *FAM13A*, *LRRC34*, *IVD*, *KIF15*, *MAD1L1*, *MAPT*, *MUC5B*, *TERC* and *TERT*. Following this, enrichment analysis was performed on the genes of each cluster to investigate whether those genes were statistically overconnected (in terms of direct gene regulation) to any of the IPF-associated genes from Allen et al. (2020). If the genes that were assigned to a particular cluster were found to be overconnected to one or more of the IPF-associated genes listed above (say the exact number of overconnected IPF-associated genes is N), then a hypergeometric test was performed to approximate the statistical significance of the finding that N out of the 14 IPF-associated genes were present within the list of overconnected genes for that cluster.

None of the 14 suspected IPF susceptibility genes from Allen et al. were assigned to Cluster 1, nor were they statistically overconnected to the genes that were assigned to this cluster. *FAM13A* was one of the genes that was assigned to Cluster 2, though it did not belong to any of the top 20 significantly enriched biological pathways. Additionally, the genes in Cluster 2 were statistically overconnected to five other IPF-associated genes. These were: *AKAP13*, *DSP*, *LRRC34*, *MAPT* and *TERT*. The hypergeometric p-value was calculated to be 0.020, indicating that it is significant that five IPF-associated genes were overconnected to the genes Cluster 2 and this is more than would be expected due to random chance. None of the IPF-associated genes from Allen et al. were found in the gene list for Cluster 3, although four were found to be statistically overconnected to the genes in this cluster. These were as follows: DSP, MAD1L1, MAPT and TERT. The statistical significance of this was approximated to be P=0.008 using a hypergeometric test, again indicating that this was significantly more than would be expected under random chance.

##### Developing the gene expression-based cluster classifier

Classification is a method of supervised machine learning that uses a correctly labelled training dataset to predict which category new observations belong in.

To determine the optimal genes to include in the classifier for the IPF data, we used an iterative algorithm which performed a greedy forward search for each cluster separately to determine the optimal combination of genes to differentiate between subjects in that cluster vs all other clusters. This was done by calculating receiver operating characteristic curves for each combination of genes and selecting the combination of genes which maximised the area under the curve (AUC). In an effort to prevent the classifier from being overfit to the discovery data, a threshold was implemented to stop the algorithm once an AUC of 0.99 had been reached. Each gene was labelled as either overexpressed or underexpressed based on whether the average expression of that gene was greater in the subjects from that particular cluster compared to the average expression across all subjects.

Making predictions with the classifier was a two-stage process. First, each subject was given a classification score for each cluster. This score was calculated as the geometric mean of the overexpressed genes for that cluster minus the geometric mean of the underexpressed genes. These scores were mean centred around zero and scaled to reflect a Z-score (i.e. standard deviation equal to 1). Ideally, subjects that belonged to a certain cluster should have had a high classification Z-score for that cluster and low classification Z-scores for the other clusters.

Then, we used the classification Z-scores to fit a multinomial logistic regression model, with cluster as the independent categorical variable and the Z-scores from each cluster as the dependent variables. This model had the ability to take data from new IPF subjects and predict which cluster they were each most likely to belong in, using only expression data from the optimal genes in the classifier. Importantly, the classifier does not use absolute levels of gene expression in order to make predictions, but instead utilizes relative gene expression between subjects. This meant that the classifier could be applied to a cohort of IPF cases (from the same study) without first requiring the removal of technical effects, which allowed for the use of validation datasets that did not contain data for healthy controls.

We tested the prediction accuracy of the classifier by using it to reassign all of the IPF subjects in the discovery datasets.

##### Comparing prognostic methods using survival analysis

Kaplan-Meier plots were used to visualise the survival over time for the validation subjects in each risk group under each method. In both cases, the log-rank test was used to test the survival curves of each risk group for equality. Univariate Cox proportional-hazards models were fit to the data with risk group as the sole covariate and time-to-death as the outcome of interest. In both cases, the low-risk group was used as the reference group. The Concordance index (C-index), the equivalent of the area under the curve (AUC) for a receiver operating characteristic (ROC) curve, and the p-values from the log-rank test were used to assess which method performed best at assigning the IPF subjects to the correct risk group and therefore predicting survival.

Following this, multivariate Cox proportional-hazards models were used to assess whether the predictions made by each method were significant predictors of mortality in the validation datasets whilst adjusting for age, sex, ancestry, FVC and $DL_{CO}$. We used the likelihood ratio test and C-index to assess whether either of the two methods of risk prediction led to a significant increase in predictive ability over a Cox PH model containing only age, sex, ancestry, FVC and $DL_{CO}$.

#### Additional Tables

**TABLE E1:** Information about the transcriptomic data in the discovery datasets and the platform used in each study.

| GEO accession | GSE38958 | GSE33566 | GSE93606 |
|---|---|---|---|
| Microarray platform | Affymetrix Human Exon 1.0 ST Array | Agilent-014850 Whole Human Genome Microarray | Affymetrix Human Gene 1.1 ST Array |
| Number of gene probes | 44,280 | 32,850 | 33,297 |
| Number of unique genes | 17,256 | 12,171 | 20,254 |

**TABLE E2:** The significantly enriched (q-value <0.05) biological processes for the 769 genes assigned to Cluster 1.

| Biological process | Enrichment score | p-value | q-value |
|---|---|---|---|
| Mitochondrial ATP synthesis coupled electron transport | 7.18 | $1.0\times10^{-7}$ | $7.8\times10^{-4}$ |
| ATP synthesis coupled electron transport | 7.12 | $1.2\times10^{-7}$ | $7.8\times10^{-4}$ |
| Respiratory electron transport chain | 6.88 | $1.4\times10^{-7}$ | $7.8\times10^{-4}$ |
| Cellular respiration | 5.95 | $1.3\times10^{-6}$ | 0.005 |
| Oxidative phosphorylation | 5.84 | $4.0\times10^{-6}$ | 0.012 |
| Electron transport chain | 5.56 | $4.3\times10^{-6}$ | 0.012 |
| Homeostasis of number of cells | 5.12 | $1.1\times10^{-5}$ | 0.024 |
| Homeostatic process | 4.54 | $1.7\times10^{-5}$ | 0.032 |

**TABLE E3:** The 20 most significantly enriched (q-value <0.05) biological processes for the 839 genes assigned to Cluster 2.

| Biological process | Enrichment score | p-value | q-value |
|---|---|---|---|
| Cell activation | 12.78 | $2.2\times10^{-27}$ | $3.7\times10^{-24}$ |
| Immune system process | 11.33 | $1.7\times10^{-25}$ | $1.4\times10^{-21}$ |
| Leukocyte activation | 11.76 | $2.4\times10^{-23}$ | $1.2\times10^{-19}$ |
| Immune response | 9.83 | $6.0\times10^{-19}$ | $2.5\times10^{-15}$ |
| Regulation of immune system process | 9.75 | $1.5\times10^{-18}$ | $4.9\times10^{-15}$ |
| Regulated exocytosis | 8.90 | $2.5\times10^{-14}$ | $6.9\times10^{-11}$ |
| Response to stimulus | 7.30 | $1.3\times10^{-13}$ | $3.1\times10^{-10}$ |
| Defence response | 8.16 | $1.6\times10^{-13}$ | $3.2\times10^{-10}$ |
| Multi-organism process | 7.74 | $1.9\times10^{-13}$ | $3.5\times10^{-10}$ |
| Lymphocyte activation | 8.73 | $4.5\times10^{-13}$ | $7.5\times10^{-10}$ |
| Translational initiation | 9.72 | $6.4\times10^{-13}$ | $9.1\times10^{-10}$ |
| Symbiotic process | 8.24 | $6.6\times10^{-13}$ | $9.1\times10^{-10}$ |
| Interspecies interaction between organisms | 8.02 | $1.6\times10^{-12}$ | $2.1\times10^{-9}$ |
| Peptide metabolic process | 8.31 | $1.9\times10^{-12}$ | $2.1\times10^{-9}$ |
| Exocytosis | 8.06 | $1.9\times10^{-12}$ | $2.1\times10^{-9}$ |
| Peptide biosynthetic process | 8.43 | $2.9\times10^{-12}$ | $2.9\times10^{-9}$ |
| Translation | 8.46 | $3.2\times10^{-12}$ | $3.1\times10^{-9}$ |
| Regulation of biological quality | 7.14 | $3.8\times10^{-12}$ | $3.5\times10^{-9}$ |
| Myeloid leukocyte activation | 8.09 | $4.1\times10^{-12}$ | $3.6\times10^{-9}$ |
| Regulation of multicellular organismal process | 7.20 | $5.0\times10^{-12}$ | $4.0\times10^{-9}$ |

**TABLE E4:** The 20 most significantly enriched (q-value <0.05) biological processes for the 784 genes assigned to Cluster 3.

| **Biological process** | **Enrichment score** | **p-value** | **q-value** |
|---|---|---|---|
| Cell activation | 20.78 | $1.3\times10^{-60}$ | $1.5\times10^{-56}$ |
| Immune response | 19.53 | $1.8\times10^{-60}$ | $1.5\times10^{-56}$ |
| Leukocyte activation | 20.87 | $3.3\times10^{-59}$ | $1.8\times10^{-55}$ |
| Immune system process | 18.04 | $1.6\times10^{-57}$ | $6.6\times10^{-54}$ |
| Immune effector process | 19.19 | $1.2\times10^{-52}$ | $4.0\times10^{-49}$ |
| Myeloid leukocyte activation | 20.63 | $1.7\times10^{-52}$ | $4.7\times10^{-49}$ |
| Leukocyte activation involved in immune response | 20.07 | $9.2\times10^{-51}$ | $2.2\times10^{-47}$ |
| Cell activation involved in immune response | 19.98 | $1.9\times10^{-50}$ | $3.9\times10^{-47}$ |
| Neutrophil activation | 20.19 | $1.0\times10^{-48}$ | $1.9\times10^{-45}$ |
| Granulocyte activation | 20.02 | $3.5\times10^{-48}$ | $5.7\times10^{-45}$ |
| Neutrophil activation involved in immune response | 19.55 | $4.0\times10^{-46}$ | $6.1\times10^{-43}$ |
| Leukocyte degranulation | 19.42 | $5.0\times10^{-46}$ | $6.8\times10^{-43}$ |
| Neutrophil degranulation | 19.43 | $1.3\times10^{-45}$ | $1.7\times10^{-42}$ |
| Myeloid cell activation involved in immune response | 19.21 | $1.5\times10^{-45}$ | $1.8\times10^{-42}$ |
| Neutrophil mediated immunity | 19.23 | $3.6\times10^{-45}$ | $3.9\times10^{-42}$ |
| Myeloid leukocyte mediated immunity | 18.99 | $1.1\times10^{-44}$ | $1.1\times10^{-41}$ |
| Leukocyte mediated immunity | 17.11 | $4.3\times10^{-43}$ | $4.2\times10^{-40}$ |
| Secretion by cell | 16.63 | $3.9\times10^{-41}$ | $3.5\times10^{-38}$ |
| Export from cell | 16.50 | $5.9\times10^{-41}$ | $5.2\times10^{-38}$ |
| Defence response | 15.95 | $1.2\times10^{-40}$ | $1.0\times10^{-37}$ |

**TABLE E5:** The 13 genes in the optimal classifier. 'Up genes' refer to genes that were more highly expressed in the subjects for that cluster compared to the mean expression across all subjects, and 'down genes' refer to genes that were less highly expressed in the subjects in that cluster.

| **Cluster 1** | | **Cluster 2** | | **Cluster 3** | |
|---|---|---|---|---|---|
| **Up genes** | **Down genes** | **Up genes** | **Down genes** | **Up genes** | **Down genes** |
| *KCNK15* | *RPF1* | *NOP58* | | *CA4* | |
| *SORBS1* | | *PSMA5* | | *BCL2A1* | |
| *HBB* | | *RASGRP1* | | *UGCG* | |
| | | *IFI30* | | | |
| | | *HLA-DRA* | | | |
| | | *ATM* | | | |

**TABLE E6:** Coefficients of the multinomial logistic regression model fit using classification scores from the genes in the classifier. Note that Cluster 1 is the reference cluster and so the coefficients for this cluster are all zero and have been omitted.

| **Cluster** | **Intercept** | **Cluster 1 score** | **Cluster 2 score** | **Cluster 3 score** |
|---|---|---|---|---|
| Blue | 3.12 | -9.75 | 8.87 | 1.66 |
| Yellow | -16.6 | -11.92 | -3.15 | 29.42 |

**TABLE E7:** Two-way tables comparing 'true' assignment of subjects from the discovery analysis (determined using COMMUNAL with 2,500 genes) to the reassignment of these subjects using the 13-gene cluster classifier.

| | | **True cluster** | | |
|---|---|---|---|---|
| | | Cluster 1 | Cluster 2 | Cluster 3 |
| **Classifier predicted cluster** | Cluster 1 | 63 | 1 | 0 |
| | Cluster 2 | 1 | 94 | 0 |
| | Cluster3 | 0 | 0 | 37 |

**TABLE E8:** Pairwise comparisons showing the differences in survival over time between any two validation clusters, estimated using Cox proportional hazards models.

| **Reference cluster** | **Alternate cluster** | **Hazard Ratio** | **95% CI** | **P-value** |
|---|---|---|---|---|
| Cluster 2 | Cluster 1 | 3.80 | 1.78, 8.12 | 0.001 |
| Cluster 2 | Cluster 3 | 5.05 | 2.24, 11.35 | $9.1\times10^{-5}$ |
| Cluster 1 | Cluster 3 | 1.47 | 0.67, 3.22 | 0.341 |

**TABLE E9:** The agreement between the cluster classifier and SAMS when validation subjects were assigned to risk groups using each method.

| **GSE27957 (n=45)** | | Cluster classifier | |
|---|---|---|---|
| | | High risk | Low risk |
| SAMS | High risk | 13 | 2 |
| | Low risk | 5 | 25 |
| **GSE28042 (n=75)** | | Cluster classifier | |
| | | High risk | Low risk |
| SAMS | High risk | 17 | 12 |
| | Low risk | 19 | 27 |
| **Both datasets combined (n=120)** | | Cluster classifier | |
| | | High risk | Low risk |
| SAMS | High risk | 30 | 14 |
| | Low risk | 24 | 52 |

**TABLE E10:** Summary statistics from the Cox proportional hazards model adjusting for cluster, age, sex, ancestry, predicted forced vital capacity (FVC) and predicted diffusing capacity of the lung for carbon monoxide ($DL_{CO}$). OR = odds ratio, SE = standard error and CI = confidence interval.

| Variable | OR | SE | P-value | 95% CI |
|---|---|---|---|---|
| Cluster (high-risk cluster) | 2.697 | 0.367 | 0.007 | (1.315, 5.534) |
| Age (years) | 1.006 | 0.020 | 0.748 | (0.968, 1.046) |
| Sex (male) | 5.720 | 0.752 | 0.020 | (1.310, 24.969) |
| Ancestry (non-European) | 1.099 | 0.608 | 0.876 | (0.334, 3.619) |
| Predicted FVC | 0.996 | 0.013 | 0.745 | (0.971, 1.022) |
| Predicted $DL_{CO}$ | 0.967 | 0.013 | 0.008 | (0.944, 0.991) |

### Additional Figures

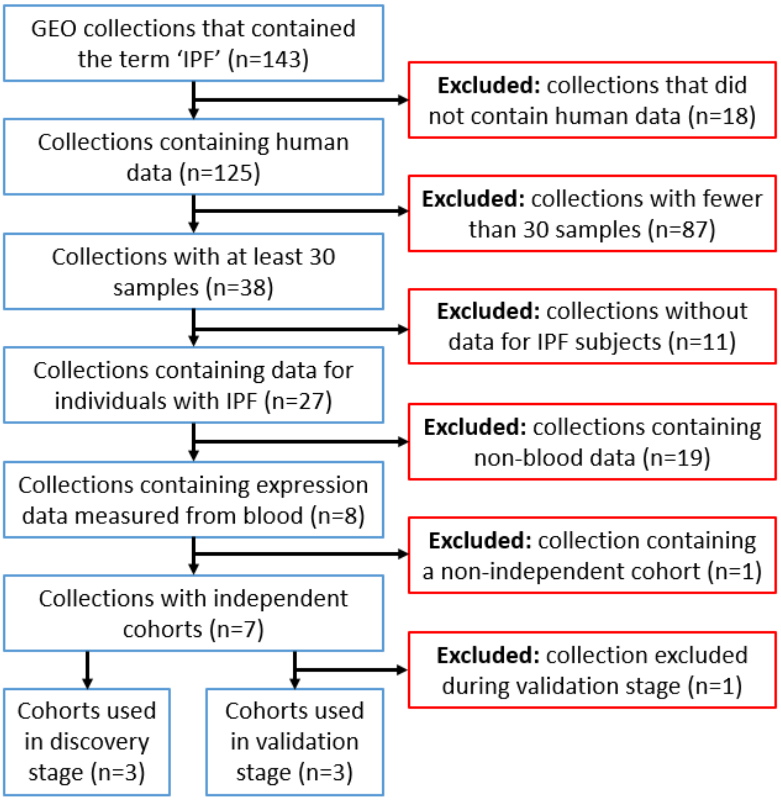

**FIGURE E1:** Flow diagram showing the process used to systematically select publicly available IPF gene expression datasets from the Gene Expression Omnibus for use in this study.

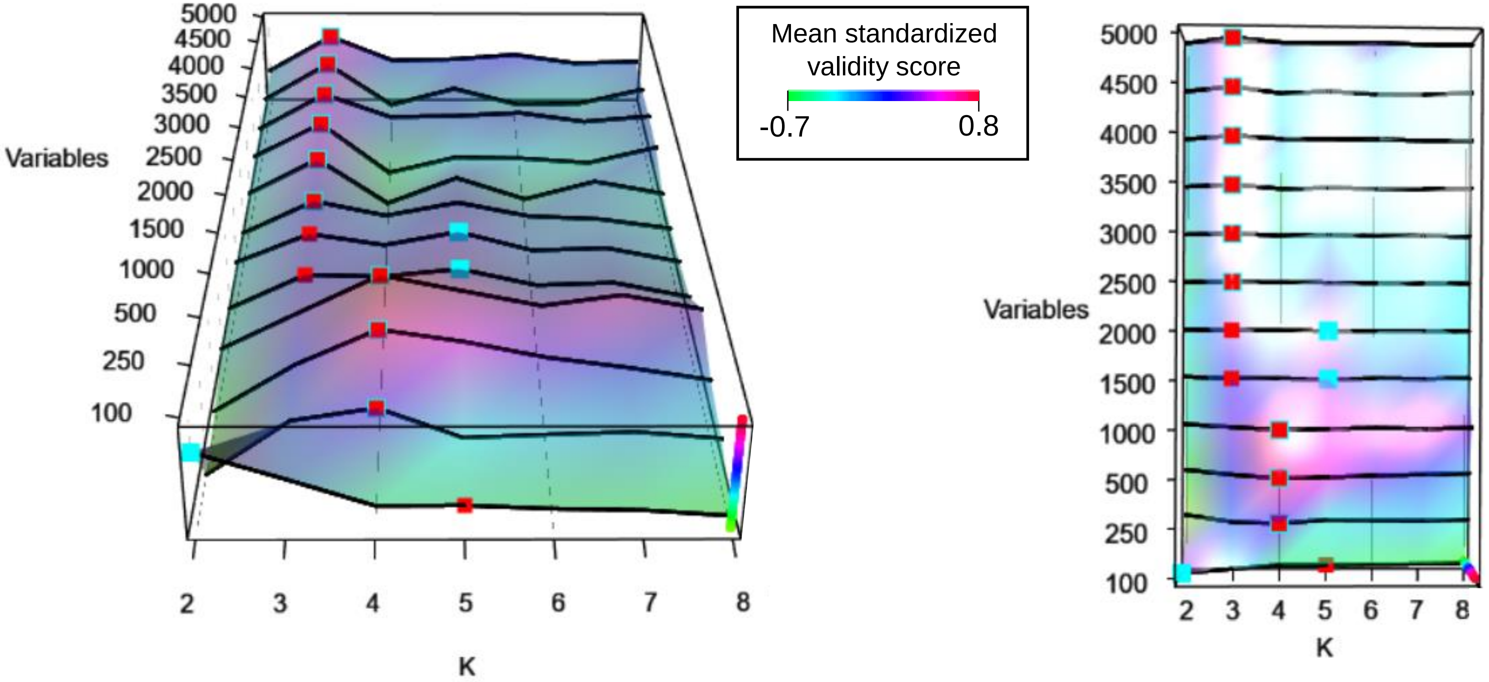

**FIGURE E2:** The 3D optimality map produced by COMMUNAL to identify the most robust number of clusters in the co-normalised data. A higher validity score indicates a better clustering assignment and stable optima are the points where the blue and red squares meet. In this map there are stable optima at K=4 from 250 genes to 1,000 genes, and at K=3 from 2,500 genes to 5,000 genes, as shown by the red and blue squares meeting. Despite the K=4 clustering assignment at 1,000 genes showing the highest mean standardized validity score of all tested clustering assignments, there were stable optima at K=3 clusters over a larger range of tested space, indicating a stronger biological signal. As such, K=3 was chosen as the optimal number of clusters in the pooled IPF dataset. The clustering at 2,500 genes and 3 clusters was chosen as the optimal clustering assignment, under the assumption that using the fewest number of genes has the least amount of redundant signal.

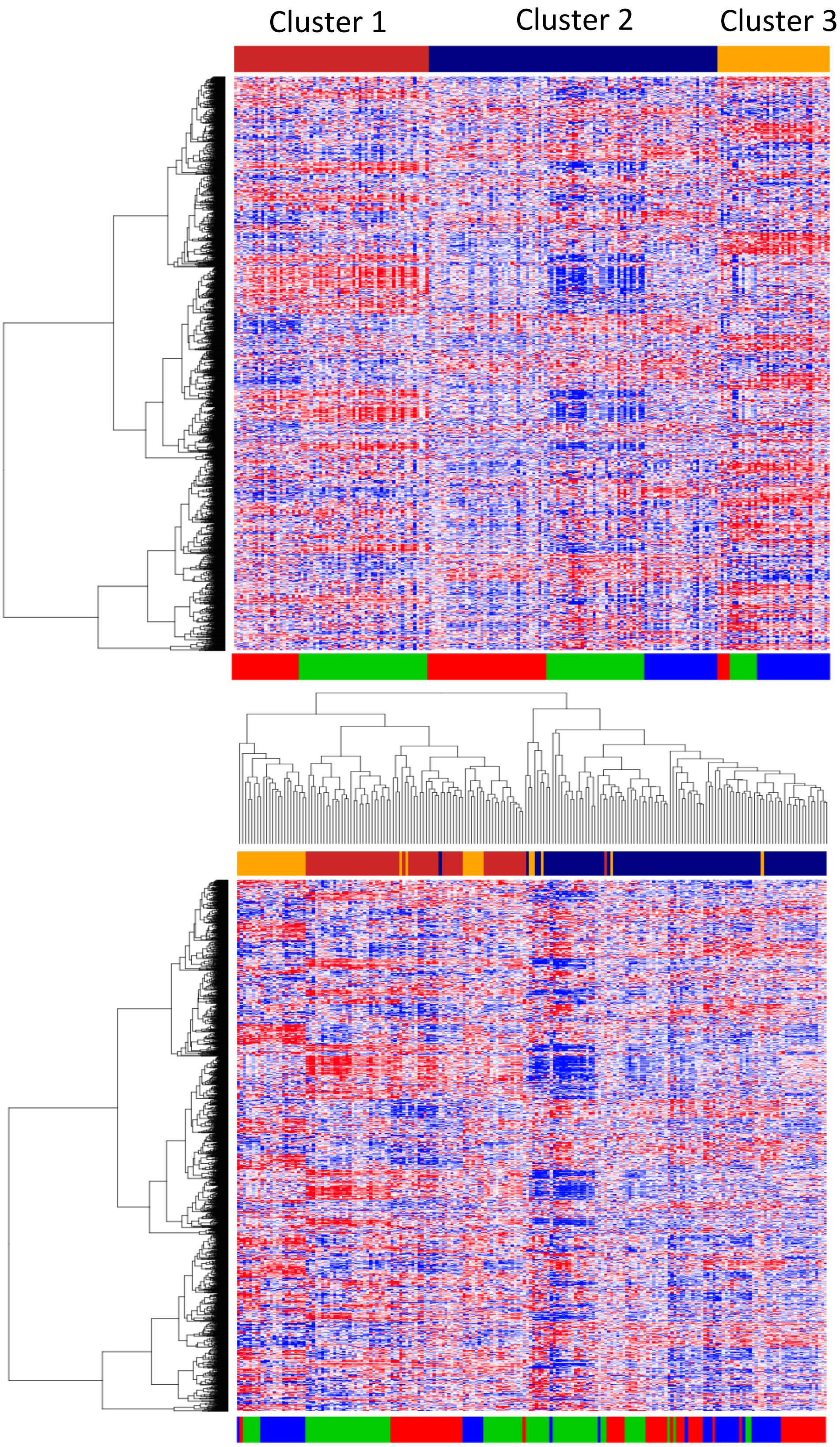

**FIGURE E3:** Heatmaps of gene expression for the clustered samples (x-axis) across the top 2,500 genes (y-axis), without hierarchical clustering of the samples (A) and with hierarchical clustering of the samples (B). Blue inside the heatmap indicates low expression and red indicates high expression. In both plots, the genes have been hierarchically clustered for presentation purposes, the bar above the plot shows the cluster that subject was assigned in to (red = cluster 1, blue = cluster 2 and yellow = cluster 3) and the bar below the plot indicates which original study the subject was in (red = GSE38958, green = GSE33566 and blue = GSE93606).

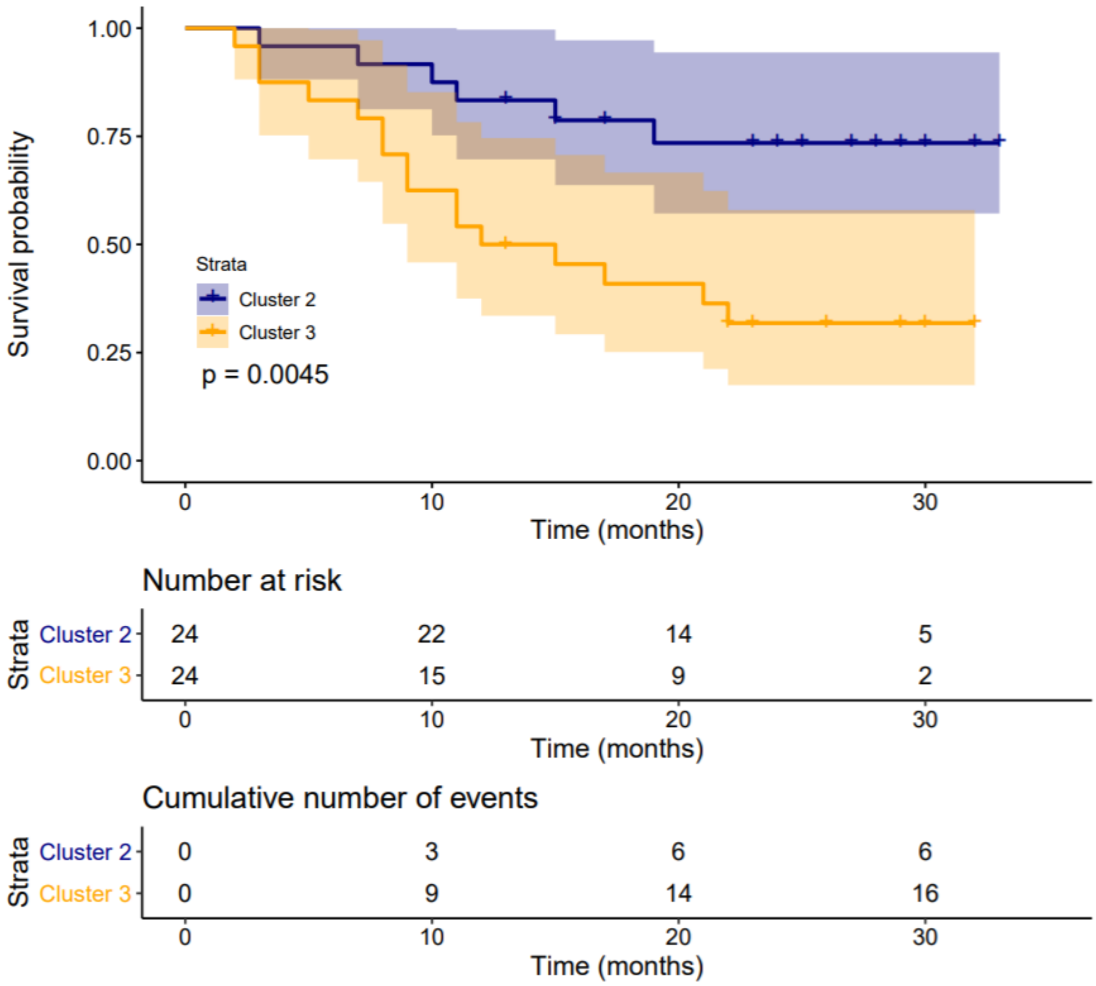

**FIGURE E4:** Kaplan-Meier curves and corresponding 95% confidence intervals showing survival over time for the subjects from study GSE93606, stratified by the cluster which they were assigned to in this study. The p-value shown on the plot is from a log-rank test testing the two curves for equality.

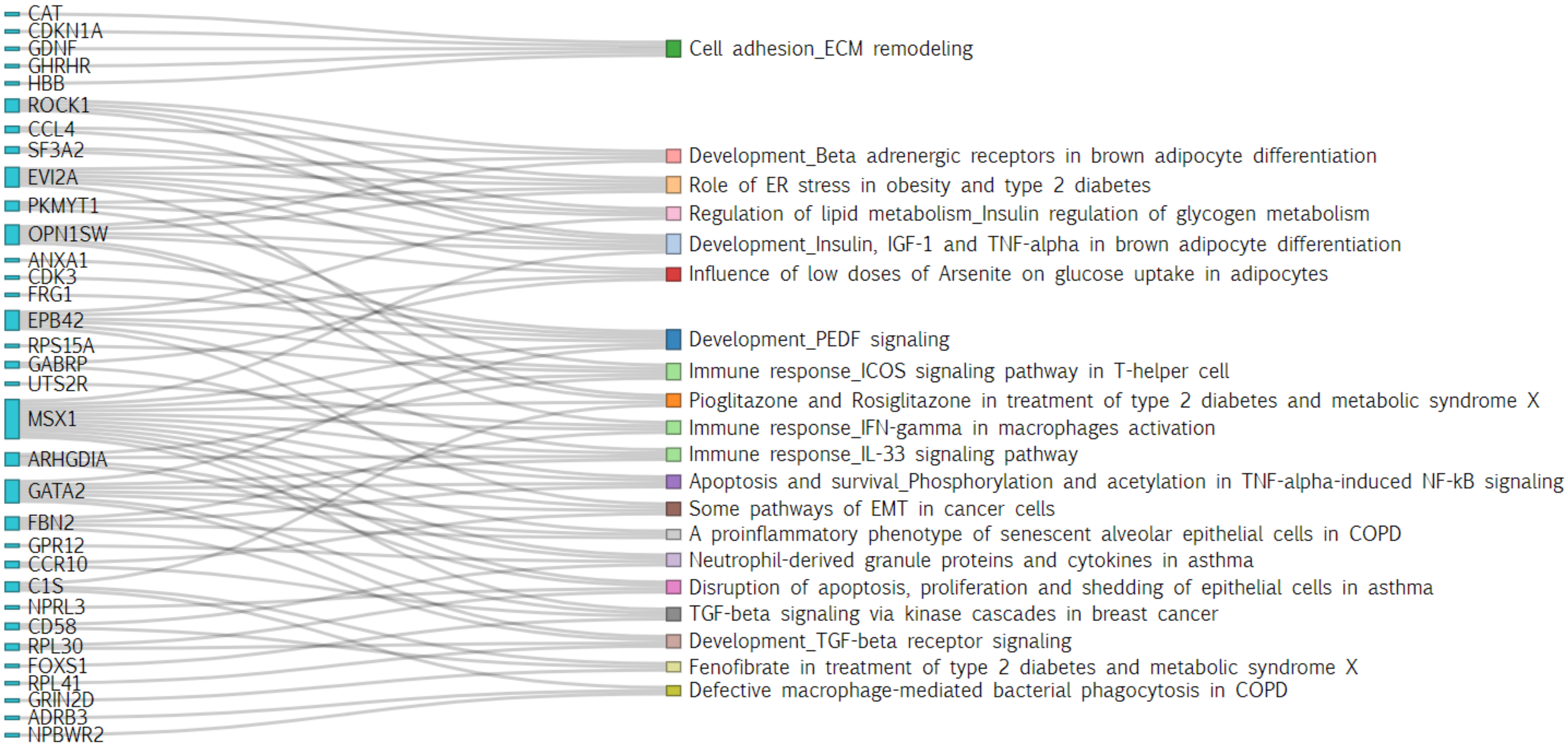

**FIGURE E5:** A Sankey diagram for Cluster 1 showing the genes that correspond to the 20 most significantly enriched biological pathways. The colour on the right hand side of the plot indicates the category of a particular pathway.

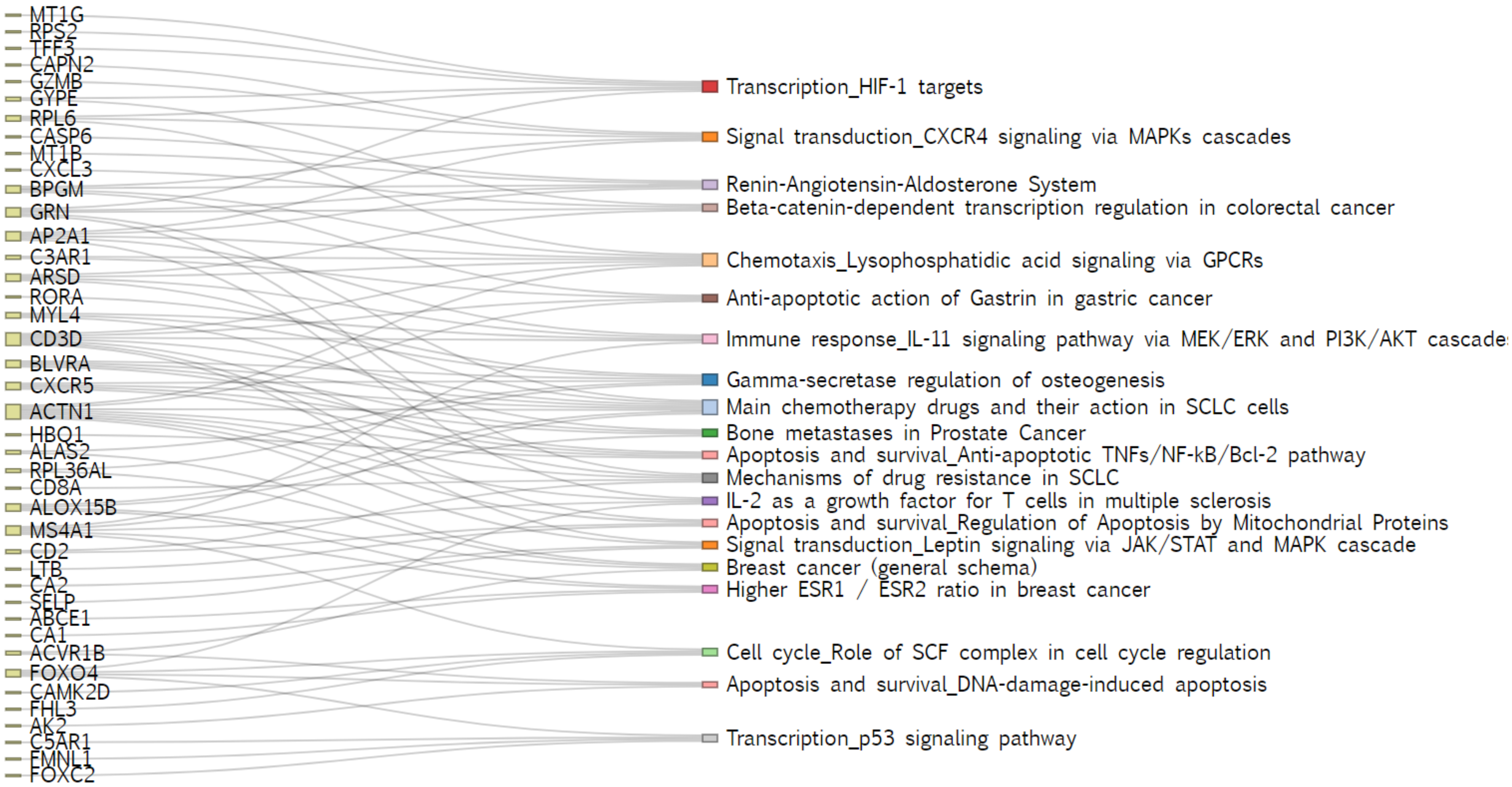

**FIGURE E6:** A Sankey diagram for Cluster 2 showing the genes that correspond to the 20 most significantly enriched biological pathways. The colour on the right hand side of the plot indicates the category of a particular pathway.

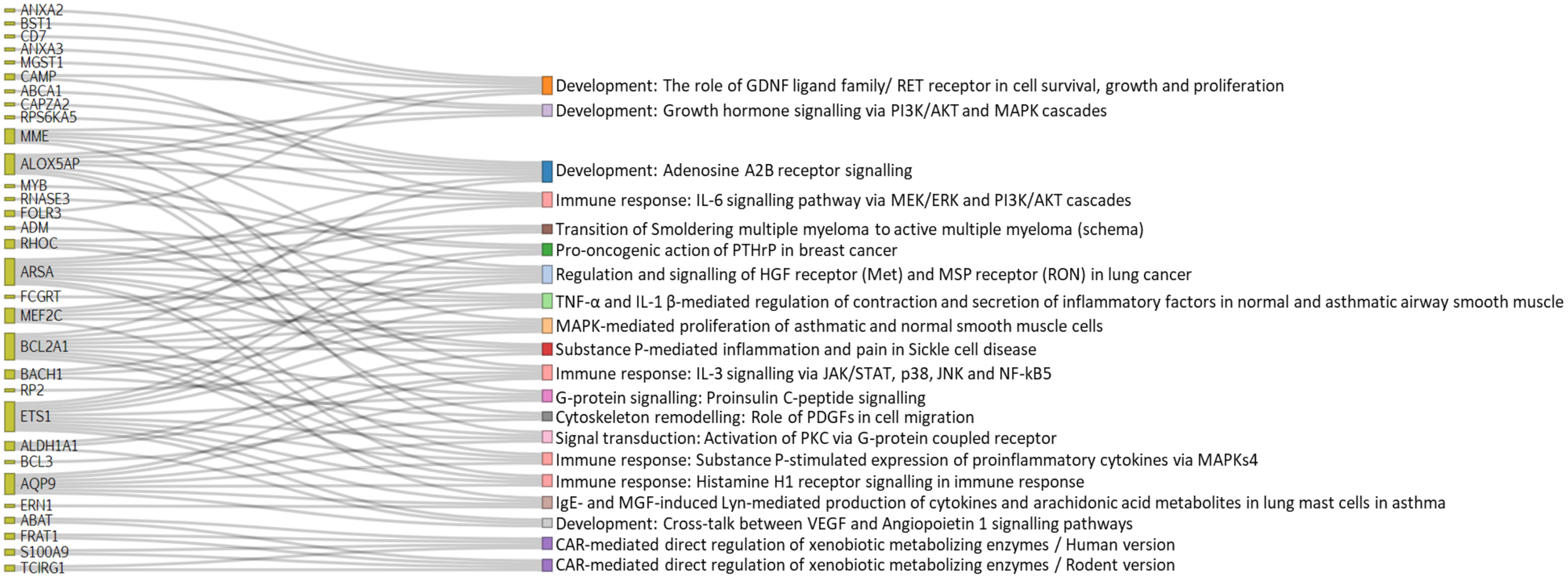

**FIGURE E7:** A Sankey diagram for Cluster 3 showing the genes that correspond to the 20 most significantly enriched biological pathways. The colour on the right hand side of the plot indicates the category of a particular pathway.

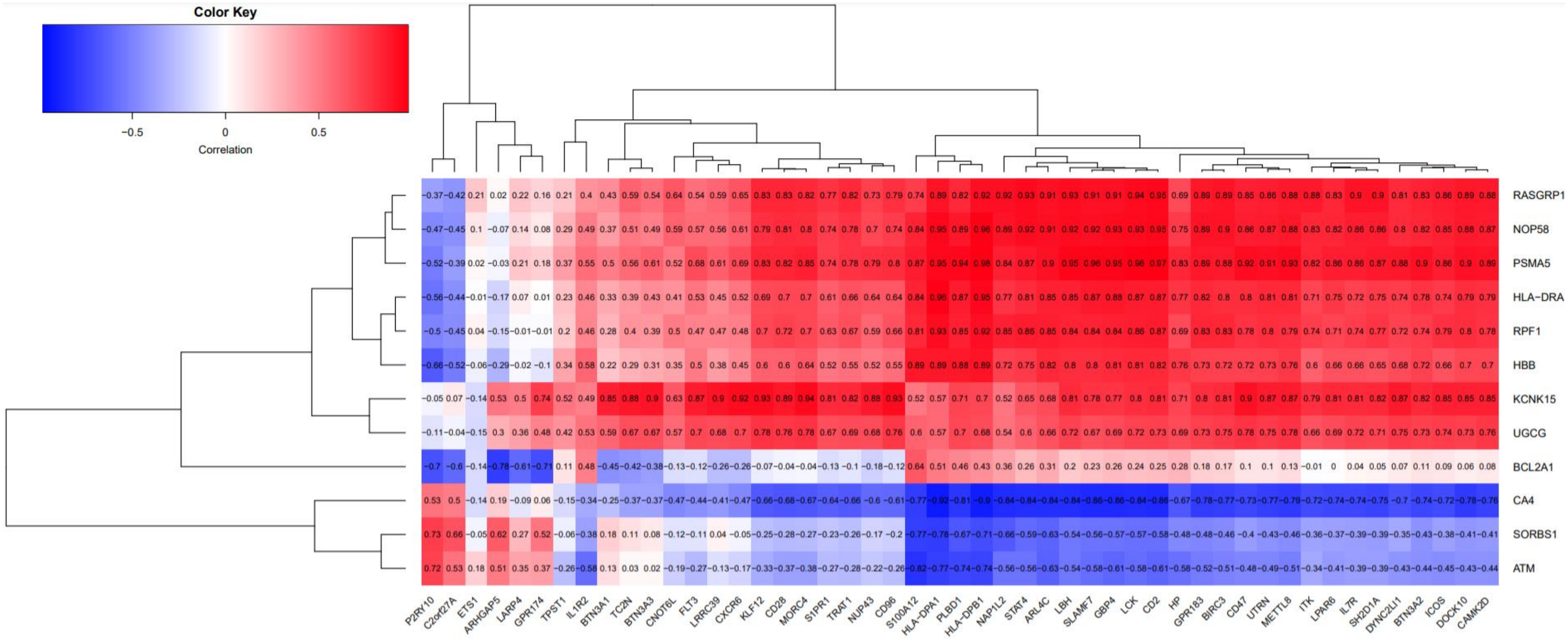

**FIGURE E8:** A heatmap showing the Pearson correlation between the genes in the classifier (x-axis) and the genes used by SAMS (y-axis). The correlation was calculated using the data from the IPF patients in the three validation cohorts (total n=194) for all genes that had complete data (12/13 genes for the classifier and 49/52 genes for SAMS). Both sets of genes were clustered using hierarchical clustering for presentation purposes.
